# Neutralization of SARS-CoV-2 Omicron variant by sera from BNT162b2 or Coronavac vaccine recipients

**DOI:** 10.1101/2021.12.13.21267668

**Authors:** Lu Lu, Bobo Wing-Yee Mok, Linlei Chen, Jacky Man-Chun Chan, Owen Tak-Yin Tsang, Bosco Hoi-Shiu Lam, Vivien Wai-Man Chuang, Allen Wing-Ho Chu, Wan-Mui Chan, Jonathan Daniel Ip, Brian Pui-Chun Chan, Ruiqi Zhang, Cyril Chik-Yan Yip, Vincent Chi-Chung Cheng, Kwok-Hung Chan, Ivan Fan-Ngai Hung, Kwok-Yung Yuen, Honglin Chen, Kelvin Kai-Wang To

## Abstract

**Background:** The SARS-CoV-2 Omicron variant, designated as a Variant of Concern(VOC) by the World Health Organization, carries numerous spike protein mutations which have been found to evade neutralizing antibodies elicited by COVID-19 vaccines. The susceptibility of Omicron variant by vaccine-induced neutralizing antibodies are urgently needed for risk assessment.

**Methods:** Omicron variant strains HKU691 and HKU344-R346K were isolated from patients using TMPRSS2-overexpressing VeroE6 cells. Whole genome sequence was determined using nanopore sequencing. Neutralization susceptibility of ancestral lineage A virus and the Omicron, Delta and Beta variants to sera from 25 BNT162b2 and 25 Coronavac vaccine recipients was determined using a live virus microneutralization assay.

**Results:** The Omicron variant strain HKU344-R346K has an additional spike R346K mutation, which is present in 8.5% of strains in GISAID database. Only 20% and 24% of BNT162b2 recipients had detectable neutralizing antibody against the Omicron variant HKU691 and HKU344-R346K, respectively, while none of the Coronavac recipients had detectable neutralizing antibody titer against either Omicron isolates. For BNT162b2 recipients, the geometric mean neutralization antibody titers(GMT) of the Omicron variant isolates(5.43 and 6.42) were 35.7-39.9-fold lower than that of the ancestral virus(229.4), and the GMT of both omicron isolates were significantly lower than those of the beta and delta variants. There was no significant difference in the GMT between HKU691 and HKU344-R346K.

**Conclusions:** Omicron variant escapes neutralizing antibodies elicited by BNT162b2 or CoronaVac. The additional R346K mutation did not affect the neutralization susceptibility. Our data suggest that the Omicron variant may be associated with lower COVID-19 vaccine effectiveness.

## INTRODUCTION

The severe acute respiratory syndrome coronavirus 2 (SARS-CoV-2) transmits efficiently among the human population [1]. Airborne transmission of SARS-CoV-2 within hospitals have been reported [2]. Stringent public health and social measures have been implemented worldwide in an effort to control the Coronavirus Disease 2019 (COVID-19) pandemic [3].

The availability of COVID-19 vaccine since late 2020 has offered hope that the pandemic can be brought under control. At the time of writing, 8 COVID-19 vaccines have been validated for use by the WHO under the Emergency Use Listing, including the inactivated whole-virion vaccine (Coronavac, BBIBP-CorV, and Covaxin), mRNA vaccine (BNT162b2 and mRNA-1273), and adenovirus-vector vaccine (Ad26.COV2.S, AZD1222, Covishield) [4]. However, several variants with mutations in the spike protein receptor binding domain (RBD), especially at the receptor binding motif (RBM), have emerged and have been found to escape neutralization from antibodies in patients recovered from COVID-19 infection or vaccine recipients [5,6]. These variants, especially the Beta, Gamma and Delta variants, have been associated with reduced vaccine effectiveness [7].

On 24^th^ November 2021, South Africa and Botswana have alerted the World Health Organization of a novel variant B.1.1.529, which has over 50 amino acid mutations, deletions or insertions when compared with the ancestral virus strains first detected in December 2019. On 26^th^ November, 2021, the World Health Organization has classified B.1.1.529 as a Variant of Concern (VOC), and has named it the Omicron variant. The B.1.1.529 lineage has been subsequently subdivided into BA.1 and BA.2. The Omicron variant appears to have transmitted via the airborne route between two travelers who were quarantined at a hotel in Hong Kong [8]. Preliminary data from South Africa suggest that the emergence of Omicron variant is associated with a higher risk of reinfection [9] and a faster doubling time [10]. Preliminary findings from the United Kingdom suggest that the Omicron variant is associated with a higher risk of transmission among household contacts than that of the Delta variant [11].

Because of the large number of mutations, especially at the spike RBD, there is a concern that the Omicron variant can escape antibody neutralization induced by the current generation COVID-19 vaccines. Here, we determined the neutralization susceptibility of two Omicron variant isolates to antibodies from COVID-19 vaccine recipients, and compared with the ancestral SARS-CoV-2 lineage A virus, the Beta variant, and the Delta variant.

## METHODS

### Patients

The virus strains hCoV-19/Hong Kong/HKU-691/2021 (HKU691) (GISAID accession number EPI_ISL_7138045) and hCoV-19/Hong Kong/HKU-344/2021 (HKU344-R346K) (GISAID accession number EPI_ISL_7357684) were isolated from the combined nasopharyngeal-throat swabs of two COVID-19 patients in Hong Kong. Serum specimens from recipients of BNT162b2 or Coronavac vaccines were collected in a vaccine trial conducted at two vaccination centers in Hong Kong [12]. This study was approved by the the Institutional Review Board of the University of Hong Kong/Hospital Authority Hong Kong West Cluster (UW 13-265; UW-21-214), and the Kowloon West Cluster REC (KW/EX-20-038[144-26]).

### Viral culture

Viral culture was performed in a biosafety level 3 facility as we described previously [6]. Briefly, TMPRSS2-expressing VeroE6 (VeroE6/TMPRSS2) cells (JCRB Cat#JCRB1819) were seeded with 100 μL of minimum essential medium (MEM) (Thermo Fisher Scientific) at 4 × 10^4^ cells/mL in 96-well plate and incubated at 37°C in a carbon dioxide incubator until confluence for inoculation. Each well was inoculated with 30 μL of clinical specimen. Virus-induced cytopathic effect was examined. Cultures with more than 50% virus-induced cytopathic effect was expanded to large volume and 50% tissue culture infective dose (TCID_50_) was determined.

The whole genome sequence of the culture isolates was determined using nanopore sequencing (Oxford Nanopore Technologies). Purified libraries were sequenced with the Oxford Nanopore MinION device using R9.4.1 flow cells. We described the lineage information using the Pangolin nomenclatures [13].

### Live virus microneutralization (MN) assay

Live virus MN assay was performed as we described previously [6]. Briefly, serum specimens were serially diluted in 2-folds from 1:10. Duplicates of each serum was mixed with 100 TCID50 of a lineage A virus (GISAID accession number: EPI_ISL_434571), a B.1.351 (Beta variant) (GISAID accession number: EPI_ISL_2423556), B.1.617.2 (delta variant) (GISAID accession number: EPI_ISL_3221329) virus isolates for 1 hour, and the serum-virus mixture was then added to VeroE6/TMPRSS2 cells. After incubation for 3 days, cytopathic effect was examined.

### Statistical analysis

Statistical analysis was performed using SPSS 26.0 (IBM SPSS Statistics) and GraphPad PRISM 9.1.1 (GraphPad Software, San Diego CA, USA). Log-transformed microneutralization antibody titers against different variants were compared using one-way ANOVA with Dunn’s multiple comparisons test. For the purpose of statistical analysis, an MN titer of <10 was considered as 5. A P value of <0.05 was considered statistically significant.

## RESULTS

This study included serum specimens from a total of 50 adult vaccine recipients, among which 25 received BNT162b2 and 25 received the Coronavac. The median age was 48 years, with an interquartile range of 40 to 54 years (Table 1). There was no statistical significant difference in the age or sex between the BNT162b2 and Coronavac groups. All individuals have received two doses of COVID-19 vaccines, and the serum specimens were collected 56 days after the first dose of vaccine.

**Table 1.** Demographics of vaccine recipients

|  | **All vaccinees**<br>(n=50) | **BNT162b2**<br>(n=25) | **CoronaVac**<br>(n=25) | **P value** |
| --- | --- | --- | --- | --- |
| **Female sex** | 31 (62) | 14 (56) | 17 (68) | 0.561<sup>a</sup> |
| **Median age in years (range)** | 48 (23-61) | 47 (23-61) | 52 (23-60) | 0.183<sup>b</sup> |

Data expressed as no. (%)
<sup>a</sup> By Fisher's exact test
<sup>b</sup> By Mann-Whitney U test

We included 2 Omicron variant culture isolates (HKU691 and HKU344-R346K) in this study. Both isolates belong to the Pangolin BA.1 lineage. These two virus isolates differ by only 2 nucleotides (Table 2). HKU-344-R346K has a non-synonymous mutation G22599A, which results in R346K mutation in the spike protein. Spike R346K mutation is present in 8.5% (131/1546) of the Omicron variant sequences deposited into GISAID database as of 10^th^ December 2021 (Supplementary Table S1).

**Table 2.**
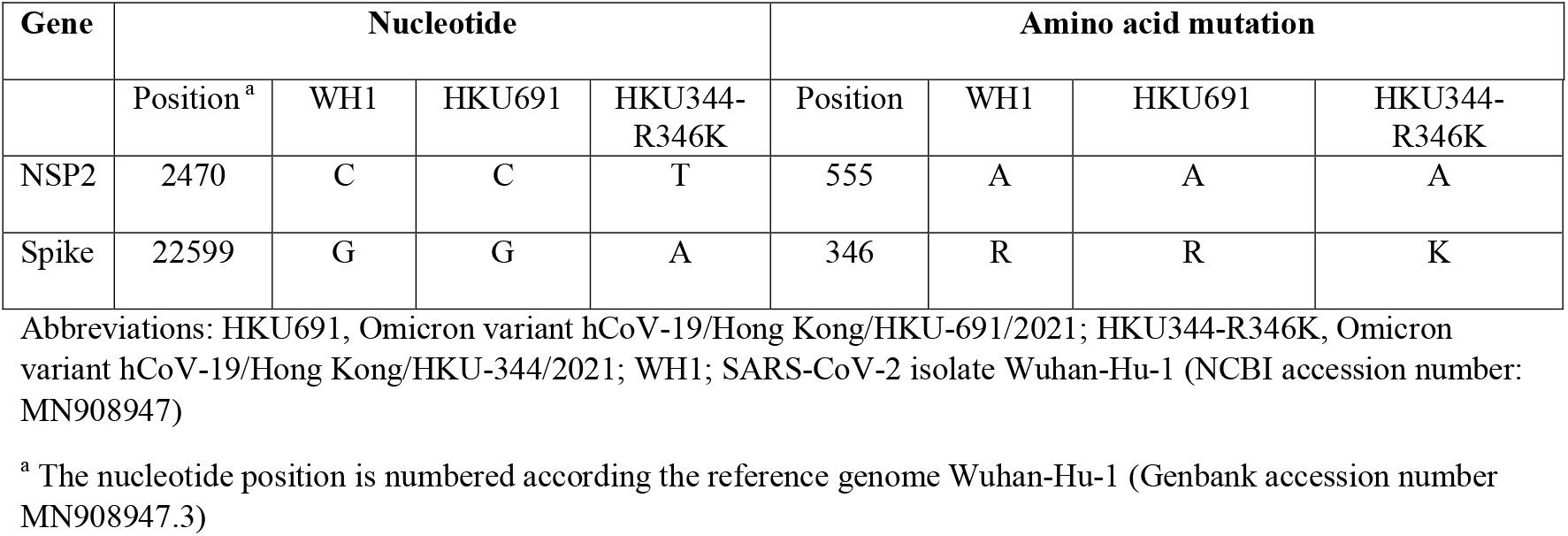
Difference in nucleotide and amino acid mutations between the two Omicron variant strains used in this study

| Gene | Nucleotide |  |  |  | Amino acid mutation |  |  |  |
| --- | --- | --- | --- | --- | --- | --- | --- | --- |
|  | Position <sup>a</sup> | WH1 | HKU691 | HKU344-R346K | Position | WH1 | HKU691 | HKU344-R346K |
| NSP2 | 2470 | C | C | T | 555 | A | A | A |
| Spike | 22599 | G | G | A | 346 | R | R | K |

Abbreviations: HKU691, Omicron variant hCoV-19/Hong Kong/HKU-691/2021; HKU344-R346K, Omicron variant hCoV-19/Hong Kong/HKU-344/2021; WH1; SARS-CoV-2 isolate Wuhan-Hu-1 (NCBI accession number: MN908947)
<sup>a</sup> The nucleotide position is numbered according the reference genome Wuhan-Hu-1 (Genbank accession number MN908947.3)

Among BNT162b2 recipients, the seropositive rate of the Omicron variants HKU691 and HKU344-R346K was only 20 and 24%, respectively, while the seropositive rates were 100% for beta variant, delta variant and ancestral virus. The geometric mean MN titer (GMT) against HKU691 (5.43) and HKU344-R346K (6.42) was 39.9 and 35.7-fold lower than those of the ancestral lineage A virus (229.4), respectively (P <0.0001) (Figure 1; Table 3). The GMT against both omicron variants were also significantly lower than those against the beta variant (25.7; P=0.0057 for HKU691 and P=0.0175 for HKU344-R346K) or the delta variant (124.7; P<0.0001). Among Coronavac recipients, none were seropositive for both Omicron variant isolates or the beta variant, while only 68% was seropositive for delta variant.

**Table 3.** Difference in GMT between Omicron variant and other earlier viruses

|  | **Lineage A**<br>**(Ancestral**<br>**virus)** | **Beta variant** | **Delta variant** | **Omicron**<br>**variant**<br>**HKU691** | **Omicron**<br>**variant**<br>**HKU344-**<br>**R346K** |
| --- | --- | --- | --- | --- | --- |
| **Seropositive**<br>**rate<sup>a</sup>**<br>**% (no.**<br>**positive/total)** |  |  |  |  |  |
| All | 100 (50/50) | 50 (25/50) | 84 (42/50) | 10 (5/50) | 12 (6/50) |
| BNT162b2 | 100 (25/25) | 100 (25/25) | 100 (25/25) | 20 (5/25) | 24 (6/25) |
| CoronaVac | 100 (25/25) | 0 (0/25) | 68 (17/25) | 0 (0/25) | 0 (0/25) |
| **Geometric**<br>**mean titer**<br>**(95% CI)** |  |  |  |  |  |
| All | 70.6<br>(48.3-103.2) | 11.3<br>(2.6-11.3) | 35.8<br>(4.0-35.8) | 5.36<br>(5.05-5.69) | 5.66<br>(5.07-6.33) |
| BNT162b2 | 229.4<br>(176.4-298.4) | 25.7<br>(19.6-33.7) | 124.7<br>(100.4-154.8) | 5.43<br>(5.11-6.46) | 6.42<br>(5.17-7.97) |
| CoronaVac | 21.7<br>(17.1-27.6) | 5<br>(5-5) | 10.3<br>(7.9-13.4) | 5<br>(5-5) | 5<br>(5-5) |
| **Fold**<br>**decrease of**<br>**GMT when**<br>**compared**<br>**with Lineage**<br>**A** |  |  |  |  |  |
| All | N/A | 6.2 | 2.0 | 13.2 | 12.5 |
| BNT162b2 | N/A | 8.9 | 1.8 | 39.9 | 35.7 |
| CoronaVac | N/A | 4.4 | 2.1 | 4.3 | 4.3 |

<sup>a</sup> MN titer $\geq 10$

**Figure 1.**
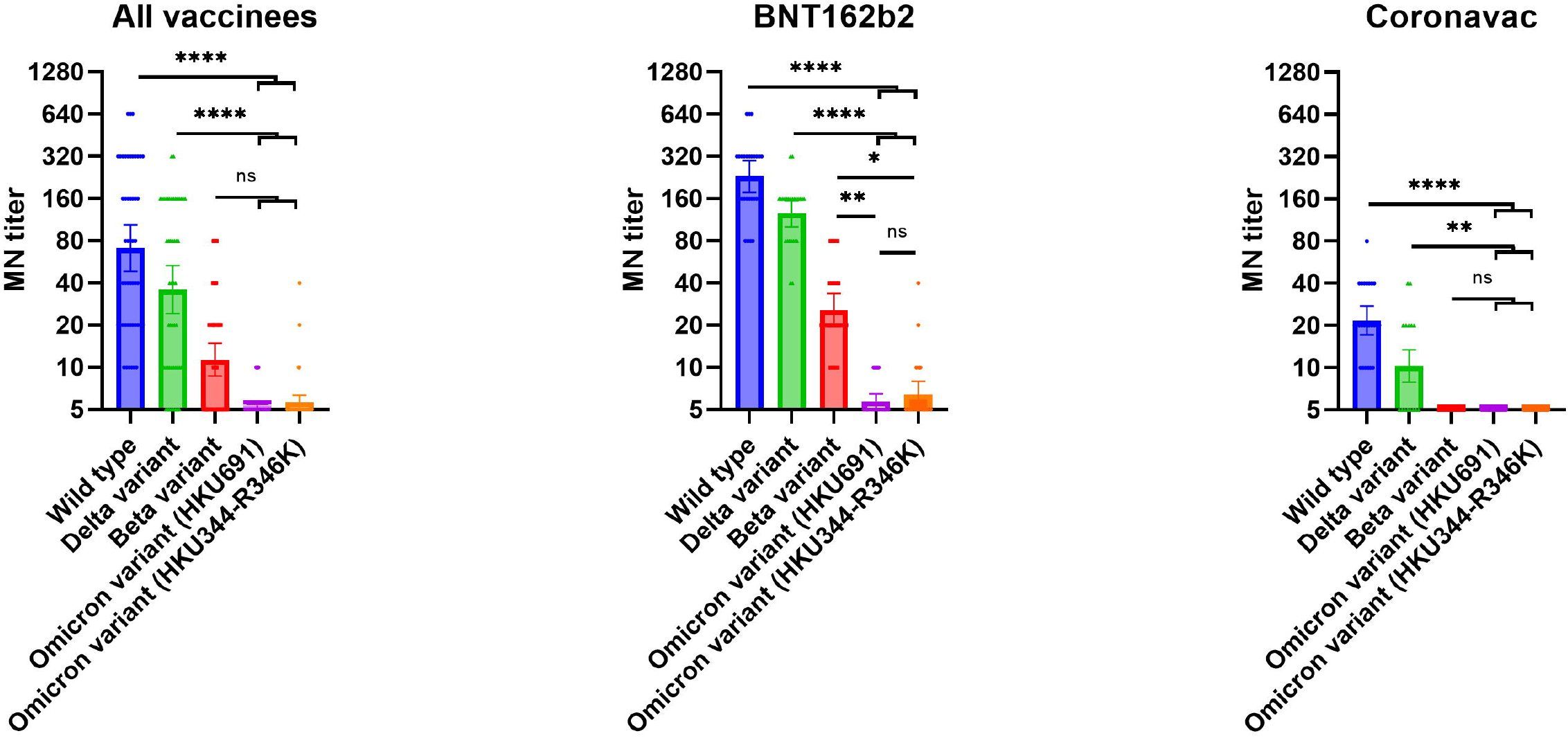
Comparison of microneutralization antibody titers between the Omicron variant and other variants or ancestral SARS-CoV-2 virus. All serum specimens were collected 56 days after the first dose of vaccine. A) Sera from all vaccine recipients. B) Sera from BNT162b2 mRNA vaccine recipients. C) Sera from Coronavac vaccine recipients. * P < .05; ** P < .01; *** P < .001; **** P < .0001.

## DISCUSSIONS

The Omicron variant poses a potential threat to vaccine effectiveness because of the presence of numerous mutations at the spike protein that have been shown to affect binding of neutralizing antibodies. In this study, we demonstrated that both Omicron variants have reduced susceptibility to neutralization by sera collected from COVID-19 vaccine recipients. None of the Coronavac recipients had detectable neutralizing antibody to the Omicron variants. The susceptibility was similar for the omicron variants with or without the spike R346K mutation.

Our results corroborate with the results from recent preprint articles. Cele et al showed that the neutralization titer is 41-fold lower than that of a D614G virus [14]. In another study, Cao et al used pseudovirus neutralization assay and yeast-display RBD library to demonstrate that the Omicron variant is shown to escape from neutralizing monoclonal antibodies [15]. Our study is unique in 2 different ways. First, we tested serum from Coronavac recipients. This is especially important because Coronavac is widely used globally. Second, we included an Omicron variant with R346K mutation. The R346K mutation is a particular concern because the spike R346K mutation is located within the RBD. R346K mutation is also present in the Mu variant, which has been shown to have to be least susceptible to sera from COVID-19 vaccine recipients [16]. The spike amino acid residue 346 is located in the epitope of a class 3 monoclonal neutralizing antibody C135 [17], and escape mutants during C135 passage contains the R346K mutation [18].

Neutralizing antibody titer correlated with vaccine protection in clinical trials and in animal studies [19,20]. Previous studies showed that the Beta (B.1.351) and Mu (B.1.621) variants are least susceptible to sera from COVID-19 vaccine recipients [5,16]. Our study showed that neutralizing antibodies generated by BNT162b2 and Coronavac vaccines are much less effective in neutralizing the Omicron variant than the Beta variant. Therefore, Omicron may be associated with a higher risk of breakthrough infection for COVID-19 vaccine recipients, or reinfection among recovered COVID-19 recipients. Indeed, preliminary data from South Africa suggest that the emergence of Omicron variant is associated with a higher risk of reinfection [9]. The Omicron variant carries important spike mutations that are present in both Beta (K417N, N501Y) and Delta (T478K) variants. The Omicron variant also contains the E484A mutation, which is located within the epitope recognized by clinically approved monoclonal antibodies bamlanivimab and casirivimab [21,22], and E484A has been shown to escape neutralization by antibody [23]. Our previous study showed that RBDs carrying E484K have lowest binding with antibodies in serum from vaccinees or from natural COVID-19 patients [5]. Furthermore, Omicron variant also carries G446S and Q493R. Mutations at spike amino acid residue 446 and 493 have been shown to affect the binding of monoclonal antibodies or from convalescent donor polyclonal serum IgG [24,25]. The effect of other mutations are less well-defined. It remains to be determined which are the critical amino acid residues that confer reduced susceptibility.

COVID-19 vaccines can also induce neutralizing antibodies that target the spike protein NTD and S2 region [26]. The Omicron variant contains several mutations in these regions. Our previous studies show that mutations in these domains can affect the neutralizing activities [6]. Whether these novel mutations affect the neutralizing activity remains to be determined.

There are several limitations in this study. First, we assessed serum specimens that were collected 56 days after the first dose of vaccine. Since antibody waning is well reported [27], it is likely that the neutralization antibody titer and seropositive rate would be lower for serum collected longer after the vaccination. Second, this study only included adult vaccine recipients. Since the antibody titer can be higher among adolescents and children, studies in the pediatric age group would be required. Third, we have not assessed the T cell immunity against the Omicron variant, which correlates with disease severity. Fourth, since all Coronavac recipients had an MN titer of <10 against the Omicron variant and the GMT against the ancestral virus is only 21.73, an accurate fold-reduction in neutralizing antibody titer cannot be determined.

In conclusion, the reduced susceptibility of Omicron variant to vaccine-induced neutralizing antibody, together with the rapid spread of this variant, suggests that newer generation vaccines should be designed to cover this variant. However, before the availability of these next generation vaccines, booster doses of currently available vaccines will likely render most people having protective levels of neutralizing antibody titers.

## Data Availability

All data produced in the present work are contained in the manuscript

## ACKNOWLEDGEMENT

We gratefully acknowledge the originating and submitting laboratories who contributed sequences to GISAID (Supplementary Table S1). This study was partly supported by the Health and Medical Research Fund, the Food and Health Bureau, The Government of the Hong Kong Special Administrative Region (HKSAR) (Ref no.: COVID1903010, Project 1), Consultancy Service for Enhancing Laboratory Surveillance of Emerging Infectious Diseases and Research Capability on Antimicrobial Resistance for Department of Health of the HKSAR; and donations of Richard Yu and Carol Yu, Shaw Foundation Hong Kong, Michael Seak-Kan Tong, May Tam Mak Mei Yin, Lee Wan Keung Charity Foundation Limited, the Providence Foundation Limited (in memory of the late Lui Hac Mih), Hong Kong Sanatorium & Hospital, Respiratory Viral Research Foundation Limited, Hui Ming, Hui Hoy and Chow Sin Lan Charity Fund Limited, Chan Yin Chuen Memorial Charitable Foundation, Marina Man-Wai Lee, the Hong Kong Hainan Commercial Association South China Microbiology Research Fund, the Jessie & George Ho Charitable Foundation, Kai Chong Tong, Tse Kam Ming Laurence, Foo Oi Foundation Limited, Betty Hing-Chu Lee, and Ping Cham So. The funding sources had no role in the study design, data collection, analysis, interpretation, or writing of the report.

