## Supplementary Table S1 for "Neutralization of SARS-CoV-2 Omicron variant by sera from BNT162b2 or Coronavac vaccine recipients"

We gratefully acknowledge the following Authors from the Originating laboratories responsible for obtaining the specimens, as well as the Submitting laboratories where the genome data were generated and shared via GISAID, on which this research is based.

All Submitters of data may be contacted directly via www.gisaid.org

Authors are sorted alphabetically.

| Accession ID | Originating Laboratory | Submitting Laboratory | Authors |
|---|---|---|---|
| EPI_ISL_7543837, EPI_ISL_7543885, EPI_ISL_7544121, EPI_ISL_7544319, EPI_ISL_7544331, EPI_ISL_7544369, EPI_ISL_7544456, EPI_ISL_7544613, EPI_ISL_7544642, EPI_ISL_7544715 | | | |
| see above | 2 Military Hospital wc MAA | NHLS/UCT | Arash Iranzadeh; Bruna Galvao; Carolyn Williamson; Deelan Doolabh; Diana Hardie; Gert Marais; Innocent Mudau; Luicer Olubayo; Lynn Tyers; Marvin Hsiao; Nokuzola Mbhele; Rageema Joseph; Stephen Korsman |
| EPI_ISL_7495248, EPI_ISL_7495249, EPI_ISL_7495250 | A. Krumbholz, Labor Dr. Krause und Kollegen MVZ GmbH, Kiel | Charité Universitätsmedizin Berlin, Institut für Virologie | Barbara Mühlemann; Christian Drosten; Julia Schneider; Julia Tesch; Jörn Beheim-Schwarzbach; Talitha Veith; Terry Jones; Tobias Bleicker; Victor M Corman |
| EPI_ISL_7405721 | A.S.L. CITTA DI TORINO - OSPEDALE AMEDEO DI SAVOIA | Fondazione del Piemonte per l'Oncologia IRCCS | Antonino Sottile; Giorgio Giardina; Paola Marino; Silvia Brossa |
| EPI_ISL_7306737 | A.S.L. TO4 | Fondazione del Piemonte per l'Oncologia IRCCS | Antonino Sottile; Giorgio Giardina; Paola Marino; Silvia Brossa |
| EPI_ISL_7016910 | ACT Pathology | Schwessinger Lab | Ashley Jones; Austin Bird; Bayantes Dagvadorj; Benjamin Schwessinger; Carolina Correa Ospina; Catalina Barragán Quintero; Craig Kennedy; Elise Kellett; Emma Crean; Evie Hodgson; Gabrielle Smith; Karina Kennedy; Rachel Leonard; Rene Riedelbauch; Robyn Hall; Salome Wilson; Scott Ferguson |
| EPI_ISL_6891760, EPI_ISL_7129657, EPI_ISL_7129884, EPI_ISL_7130085, EPI_ISL_7130189, EPI_ISL_7130344, EPI_ISL_7130486 | | | |
| see above | AGES-Institute for medical Microbiology and Hygiene Vienna | AGES-Institute for medical Microbiology and Hygiene Vienna | Alexander Indra; Elisabeth Polster; Florian Heger; Julia Klikovits; Kathrin Lippert; Marion Blaschitz; Patrick Hyden; Peter Hufnagl; Stefanie Dobrovolny; Vera Wallner |
| EPI_ISL_7381191 | AHRI | CERI, Centre for Epidemic Response and Innvoation, Stellenbosch University and KRISP, KZN Research Innovation and Sequencing Platform, UKZN. | Arisha Maharaj; Bernstein Mallory; Cele Sandile; Giandhari J; Karim Farina; Khan Khadija; Moir M; Naidoo Y; Pillay S; Ramphal U; Ramphal Y; San JE; Sigal Alex; Tegally H; Tshiabuila D; Wilkinson E; de Oliveira T; van Wyk S |
| EPI_ISL_7358094 | AHRI-Sigal | CERI, Centre for Epidemic Response and Innvoation, Stellenbosch University and KRISP, KZN Research Innovation and Sequencing Platform, UKZN. | Bernstein Mallory; Cele Sandile; Giandhari J; Karim Farina; Khan Khadija; Moir M; Naidoo Y; Nokukhanya Mdlalose; Pillay S; Ramphal U; Ramphal Y; San JE; Sigal Alex; Tegally H; Tshiabuila D; Wilkinson E; de Oliveira T |
| EPI_ISL_7154405 | Aegis Sciences Corporation | Centers for Disease Control and Prevention Division of Viral Diseases, Pathogen Discovery | Alec Vest; Benjamin Rambo-Martin; Christopher Gulvick; Clinton Paden; Cyndi Clark; Dakota Howard; Dhwani Batra; Dillon Nall; Duncan MacCannell; Erisa Sula; Ethan Sanders; Holly Houdeshell; Jason Caravas; Kristine Lacek; Matthew Hardison; Matthew Schmerer; Ola Kvalvaag; Patrick Campbell; Peter Cook; Rob Case; Scott Sammons; Shatavia Morrison; Shaun Westlund; Tymeckia Kendall; Victoria Caban Figueroa; Vikramsinha Ghorpade; Yvette Unoarumhi |
| EPI_ISL_7452769, EPI_ISL_7452775 | Aesculabor Hamburg, Institut der Labormedizin | Heinrich Pette Institute, Leibniz Institute for Experimental Virology | Adam Grundhoff; Alexis Robitaille; Johannes Knobloch; Martin Aepfelbacher; Nicole Fischer; Thomas Günther |
| EPI_ISL_7470217, EPI_ISL_7470278, EPI_ISL_7470360, EPI_ISL_7470368, EPI_ISL_7470391, EPI_ISL_7470446 | Akershus University Hospital, Department for Microbiology and Infectious Disease Control | Norwegian Institute of Public Health, Department of Virology | Atiya R Ali; Debech Nadia; Engebretsen Serina Beate; Garcia Llorente Ignacio; Hilde Elshaug; Hilde Vollan; Jon Bråte; Kamilla Heddeland Instefjord; Karoline Bragstad; Kathrine Stene-Johansen; Line Victoria Moen; Marie Paulsen Madsen; Olav Hungnes; Pedersen Benedikte Nevjen; Rasmus Riis Kopperud |
| EPI_ISL_7146436, EPI_ISL_7506705 | Allergy, Immunology and Cell Biology Unit (AICBU) | Allergy, Immunology and Cell Biology Unit (AICBU) | Ayesha Wijesinghe; Chandima Jeewandara; Deshni Jayathilaka; Dinuka Ariyaratne; Diyanath Ranasinghe; Dumni Guasinghe; Farha Bary; Gathsaurie Neelika Malavige; Tibutius Thanesh |
| EPI_ISL_7544441 | Alma CDC wc AHC | NHLS/UCT | Arash Iranzadeh; Bruna Galvao; Carolyn Williamson; Deelan Doolabh; Diana Hardie; Gert Marais; Innocent Mudau; Luicer Olubayo; Lynn Tyers; Marvin Hsiao; Nokuzola Mbhele; Rageema Joseph; Stephen Korsman |
| EPI_ISL_7264139, EPI_ISL_7265455, EPI_ISL_7265456 | Alpha Labs | National Microbiology Laboratory (NML) | Anna Majer; Anneliese Landgraff; CanCOGeN's metadata curation team; Darian Hole; Dynacare Brampton COVID-19 Diagnostic team; Elsie Grudeski; Gary Van Domselaar; Gordon Jolly; Grace Seo; Jennifer Tanner; Madison Chapel; Morag Graham; Natalie Knox; Nathalie Bastien; Philip Mabon; Public Health Agency of Canada CanCOGeN team; Rhiannon Huzarewich; Russell Mandes; Shari Tyson; Timothy Booth; Yan Li |
| EPI_ISL_6914011, EPI_ISL_6914012, EPI_ISL_6914013, EPI_ISL_6914014, EPI_ISL_6914015, EPI_ISL_6914016, EPI_ISL_6914017, EPI_ISL_6914018, EPI_ISL_6914019, EPI_ISL_6914020, EPI_ISL_6914021, EPI_ISL_6914022, EPI_ISL_6914023, EPI_ISL_6914024, EPI_ISL_6914025, EPI_ISL_6914026, EPI_ISL_6914027, EPI_ISL_6914028, EPI_ISL_6914029, EPI_ISL_6914030, EPI_ISL_6914031, EPI_ISL_6914032, EPI_ISL_6914033, EPI_ISL_6914034, EPI_ISL_6914035, EPI_ISL_6914036 | | | |
| see above | Ampath Laboratories | National Institute for Communicable Diseases of the National Health Laboratory Service | Amoako DG; Bhiman JN; Everatt J; Ismail A; Mahlangu B; Mnguni A; Mohale T; Ntuli N; Scheepers C; Wolter N |
| EPI_ISL_7011321 | Area of Virology, Serology and Virology Division (SAViD), New South Wales Health Pathology Randwick | Virology Research Laboratory; Area of Virology, Serology and Virology Division (SAViD), New South Wales Health Pathology Randwick | Au, J.; Bull, R.; Deveson, I.; Foster, C.; Rawlinson, W.; Ruiz Silva, M.; Van Hal, S. |
| EPI_ISL_7503376, EPI_ISL_7503377, EPI_ISL_7503378, EPI_ISL_7548950, EPI_ISL_7548951, EPI_ISL_7548952 | Arizona State University | Arizona State University | Efrem S. Lim; Joshua LaBaer; LaRinda A. Holland; Matthew F. Smith; Nathaniel Johnson; Regan A. Sullins; Steven C. Holland; Vel Murugan |
| EPI_ISL_6963510 | Auriga Research Pvt.Ltd / Strand Life Sciences | National Centre for Biological Sciences, TIFR - Rockefeller Foundation | Aarati Karaba; Anson Kunjumon George; Aparnaa Ramanathan; Apurva Sarin; Chandrasekhar Vadlamudi; Chitra Pattabiraman; Darshan Sreenivas; Dasaradhi Palakodeti; Dimple Notani; Divya Priya A; Madhusudhan J; Manisha Bharadwaj; Manoj Kumar Jha; Mudasir Nazaar; Pradeep B P; Priyanka Ananta Mulay; Ramesh Hariharan; Rohan Pais; Satyajit Mayor; Saumitra Mardikar; Srivathsan Adimoolam; Uma Ramakrishnan; Vamsi Veeramachaneni; Vasanthapuram Ravi; Vijay Chandru; Vishal G Rao; Yasodha Kannan |
| EPI_ISL_7195246, EPI_ISL_7195247, EPI_ISL_7195248 | Austrian Agency for Health and Food Safety (AGES) | Bergthaler laboratory, CeMM Research Center for Molecular Medicine of the Austrian Academy of Sciences | Andreas Bergthaler; Anna Schedl; Bekir Erguner; Benedikt Agerer; Christoph Bock; Fabian Amman; Jan Laine; Lukas Endler; Martin Senekowitsch; Matthew Thornton; Michael Schuster; Michelle Chan; Petr Triska; Thomas Penz |
| EPI_ISL_7502111 | Ayass Bioscience LLC | Ayass Bioscience LLC | Kevin Zhu; Lina Abi Mosleh; Mohamad Ammar Ayass; Natalya Griko; Nazanin Taheri |
| EPI_ISL_7400565 | Azienda Ospedaliera Pugliese Ciaccio di Catanzaro SOC Microbiologia e Virologia | Azienda Ospedaliera Pugliese Ciaccio di Catanzaro SOC Microbiologia e Virologia | Rossana Tallerico Cinzia Peronace Federica Pasceri Marco De Fazio Ilenia Talotta Giuseppina Panduri Pasquale Minchella |
| EPI_ISL_7338921 | Azienda Sanitaria dell´Alto Adige - Laboratorio Aziendale di Microbiologia e Virologia | Azienda Sanitaria dell'Alto Adige | Irene Bianconi |
| EPI_ISL_7226262 | BNH Hospital | National Institute of Health, Department of Medical Sciences, Ministry of Public Health, Thailand | Archawin Rojanawiwat; Ballang Uppapong; Beth Skaggs; Donlaya Maunplueng; Kazuhisa Okada; Nuttida Thongpramul; Pakorn Piromtong; Pilailuk Akkapaiboon Okada; Piroon Jenjaroenpun; Pongpun Sawatwong; Prapat Suriyaphol; Sirikanda Wimol; Siripaporn Phuygun; Sittiporn Parnmen; Supakit Sirilak; Suratchana Mitrat; Thanutsapa Thanadachakul; Thidathip Wongsurawat |
| EPI_ISL_7149647, EPI_ISL_7197950, EPI_ISL_7346925, EPI_ISL_7347257, EPI_ISL_7347522, EPI_ISL_7355779, EPI_ISL_7391885, EPI_ISL_7391908, EPI_ISL_7391931, EPI_ISL_7391984, EPI_ISL_7392005, EPI_ISL_7392027, EPI_ISL_7392079, EPI_ISL_7392087, EPI_ISL_7392093, EPI_ISL_7392139, EPI_ISL_7485998, EPI_ISL_7486106, EPI_ISL_7486193, EPI_ISL_7486452, EPI_ISL_7486697, EPI_ISL_7486742, EPI_ISL_7486808, EPI_ISL_7486814, EPI_ISL_7486850, EPI_ISL_7486923, EPI_ISL_7487524, EPI_ISL_7487623, EPI_ISL_7487665, EPI_ISL_7487692, EPI_ISL_7512352, EPI_ISL_7512373, EPI_ISL_7512378, EPI_ISL_7512381, EPI_ISL_7512410, EPI_ISL_7512438, EPI_ISL_7512444, EPI_ISL_7512448, EPI_ISL_7520500, EPI_ISL_7538419, EPI_ISL_7538437, EPI_ISL_7538529, EPI_ISL_7538539, EPI_ISL_7538562, EPI_ISL_7538591, EPI_ISL_7538628, EPI_ISL_7538662, EPI_ISL_7538687, EPI_ISL_7538812, EPI_ISL_7538904, EPI_ISL_7538983, EPI_ISL_7538987, EPI_ISL_7539099, EPI_ISL_7539114, EPI_ISL_7539140, EPI_ISL_7539157, EPI_ISL_7539216, EPI_ISL_7539258, EPI_ISL_7539330, EPI_ISL_7539539, EPI_ISL_7539677, EPI_ISL_7539761 | | | |
| see above | Berkshire and Surrey Pathology Services Lighthouse Laboratory | Wellcome Sanger Institute for the COVID-19 Genomics UK (COG-UK) Consortium | Berkshire and Surrey Pathology Services Lighthouse Laboratory and Alex Alderton; Cordelia Langford; David K. Jackson; Dominic Kwiatkowski; Ewan Harrison; Ian Johnston; Jeffrey Barrett; John Sillitoe on behalf of the Wellcome Sanger Institute COVID-19 Surveillance Team; Roberto Amato; Sonia Goncalves |
| EPI_ISL_7464464, EPI_ISL_7464465 | Biolab Diagnostic Laboratories | Biolab Diagnostic Laboratories | Ahmad Tibi; Amid Abdelnour; Badia Saddedin; Eiad Atwa; Issa Abu-Dayyeh; Lama Hussein; Shaima Ali |
| EPI_ISL_7042354 | Biolytix AG | Clinical Bacteriology, University Hospital Basel | Adrian Egli; Alfredo Mari; Fanny Wegner; Hans Hirsch; Helena MB Seth-Smith; Julia Bielicki; Karoline Leuzinger; Manuel Battegay; Tim Roloff |
| EPI_ISL_7417514 | Bioscientia Labor Wermsdorf | Robert Koch Institute | |
| EPI_ISL_7544135, EPI_ISL_7544479, EPI_ISL_7544573 | Bothasig CDC wc BLD | NHLS/UCT | Arash Iranzadeh; Bruna Galvao; Carolyn Williamson; Deelan Doolabh; Diana Hardie; Gert Marais; Innocent Mudau; Luicer Olubayo; Lynn Tyers; Marvin Hsiao; Nokuzola Mbhele; Rageema Joseph; Stephen Korsman |
| EPI_ISL_7548902, EPI_ISL_7548903, EPI_ISL_7548904, EPI_ISL_7548905, EPI_ISL_7548906, EPI_ISL_7548909, EPI_ISL_7548914, EPI_ISL_7548917, EPI_ISL_7548931 | | | |

| | | | |
|---|---|---|---|
| see above | Botswana Harvard HIV Reference Laboratory | Botswana Harvard AIDS Institute Partnership, Plot 1836 North Ring Road, Princess Marina Hospital, Gaborone | Boitumelo Zuze; Botshelo Radibe; Dorcas Maruapula; Doreen Ditshwanelo; Joseph Makhema; Keoratile Ntshambiwa; Kgomotso Moruisi; Legodile Kooepile; Mosepele Mosepele; Mphaphi B. Mbulawa; Ontlametse T. Bareng; Pamela Smith-Lawrence; Roger Shapiro; Sefetogi Ramaologa; Shahin Lockman; Sikhulile Moyo; Simani Gaseitsiwe; Thongbotho Mphoyakgosi; Wonderful T. Choga |
| EPI_ISL_6640916, EPI_ISL_6640917, EPI_ISL_6640919, EPI_ISL_6670244, EPI_ISL_6752026, EPI_ISL_6752027, EPI_ISL_6774081, EPI_ISL_6774083, EPI_ISL_6774084, EPI_ISL_6774085, EPI_ISL_6774087, EPI_ISL_6774088, EPI_ISL_6774089, EPI_ISL_6774090, EPI_ISL_6774091, EPI_ISL_6774093, EPI_ISL_7121195, EPI_ISL_7121204, EPI_ISL_7121205, EPI_ISL_7380512, EPI_ISL_7380515, EPI_ISL_7380524 | | | |
| see above | Botswana Harvard HIV Reference Laboratory | Botswana Harvard HIV Reference Laboratory | Boitumelo Zuze; Botshelo Radibe; Dorcas Maruapula; Joseph Makhema; Keoratile Ntshambiwa; Kgomotso Moruisi; Legodile Kooepile; Mosepele Mosepele; Mphaphi B. Mbulawa; Ontlametse T. Bareng; Pamela Smith-Lawrence; Roger Shapiro; Sefetogi Ramaologa; Shahin Lockman; Sikhulile Moyo; Simani Gaseitsiwe; Thongbotho Mphoyakgosi; Wonderful T. Choga |
| EPI_ISL_7132804, EPI_ISL_7370157, EPI_ISL_7370181, EPI_ISL_7370259, EPI_ISL_7370472 | British Columbia Centre For Disease Control | BCCDC Public Health Laboratory | 655 W 12th Avenue; Ana Pacagnella; BC Canada V5T 2N3; Corrinne Ng; Dan Fornika; John Tyson; Kim Macdonald; Kimia Kamelian; Linda Hoang; Loretta Janz; Mel Krajden; Prystajecky Natalie; Robert Azana; Shannon Russell; Vancouver |
| EPI_ISL_7390696, EPI_ISL_7391208, EPI_ISL_7453171, EPI_ISL_7453991, EPI_ISL_7454277, EPI_ISL_7542815, EPI_ISL_7543162, EPI_ISL_7543543, EPI_ISL_7543579, EPI_ISL_7544427, EPI_ISL_7544520, EPI_ISL_7547858, EPI_ISL_7548157, EPI_ISL_7550313, EPI_ISL_7550474, EPI_ISL_7550515, EPI_ISL_7550529, EPI_ISL_7550748 | | | |
| see above | Broad Institute Clinical Research Sequencing Platform | Infectious Disease Program, Broad Institute of Harvard and MIT | Adams, G.; B.L.; B.W.; Bauer, M.; Birren; Blumenstiel, B.; Brown, C.; Carter, A.; Chaluvadi, S.; D.J.; DeFelice, M.; DeRuff, K.; Dodge, S.; Gabriel, S.; Gallagher, G.; Gladden-Young, A.; Granger, B.; J.E.; K.J.; Lagerborg, K.; Larkin, K.; Lee, M.; Lemieux; Lennon, N.; Loreth, C.; Madoff, L.; McGovern, S.; Meldrim, J.; Normandin, E.; P.C.; Park; Pearlman, L.; Reilly, S.; Rudy, M.; Sabeti; Siddle; Smole, S.; Tomkins-Tinch, C.; Vicente, G.; and MacInnis |
| EPI_ISL_7263830 | CAP Roger de Flor | Banc de Sang i Teixits | Carlos Hobeich; Francisco Vidal; Irene Corrales; Lorena Ramírez; Maria Glòria Soria; Natália Comes; Nina Borràs; Noemí Gonzalez; Silvia Sauleda |
| EPI_ISL_7477252 | CDPH VBL | California Department of Public Health | Emily Smith on behalf of CDPH-COVIDNet |
| EPI_ISL_7427856 | CENTOGENE Frankfurt Laboratory: Niederlassung Industriepark Höchst | Robert Koch Institute | |
| EPI_ISL_7547731 | CENTOLAB | National Reference Laboratory, Nigeria Centre for Disease Control | Catherine Okoi; Chimaobi Chukwu; Dr Ifedayo Adetifa; Dr Ndodo Nnaemeka; Dr Omoare Adesuyi; Nwando Mba; Olajumoke Babatunde; Olusola Anuoluwapo Akanbi; Oyeronke Ayansola |
| EPI_ISL_6862897 | CERBALLIANCE | UMR PIMIT | David A Wilkinson; Patrick Mavingui |
| EPI_ISL_6962948, EPI_ISL_6967758 | CERBALLIANCE PARIS ET IDF EST | CERBA HealthCare | Bénédicte Roquebert; Johanna Roux; Judith Zerah; Laura Verdurme; Sabine Trombert; Stéphanie Haim-Boukobza |
| EPI_ISL_7226961 | CHU de Bordeaux | CNR Virus des Infections Respiratoires - France SUD | Antonin Bal; Bruno Lina; Bruno Simon; Denis Malvy; Gregory Destras; Gwendolyne Burfin; Hadrien Regue; Laurence Josset; Marie-Edith Lafon; Martine Valette; Pantxika Bellecave; Quentin Semanas |
| EPI_ISL_7268670, EPI_ISL_7269933 | CLINA-LANCET LABORATORIES | National Reference Laboratory, Nigeria Centre for Disease Control | Catherine Okoi; Chimaobi Chukwu; Dr Ifedayo Adetifa; Dr Ndodo Nnaemeka; Dr Omoare Adesuyi; Nwando Mba; Olajumoke Babatunde; Olusola Anuoluwapo Akanbi; Oyeronke Ayansola |
| EPI_ISL_7313633 | CT Department of Public Health | CT Department of Public Health | Claire Pearson; Tu N. Nguyen |
| EPI_ISL_7137326, EPI_ISL_7137327, EPI_ISL_7137328, EPI_ISL_7137330 | California Department of Public Health | California Department of Public Health | CDPH IDLB COVIDNet |
| EPI_ISL_7217437, EPI_ISL_7217563 | Cantacuzino National Military-Medical Institute, Viral Respiratory Infections Laboratory | Cantacuzino Institute Virology | Luiza Ustea; Mihaela Lazar; Mihaela Oprea; Nicoleta Paraschiv; Sorin Dinu |
| EPI_ISL_6980876 | Cerballiance, Reunion | UMR PIMIT | David A Wilkinson; Patrick Mavingui |
| EPI_ISL_7019047 | Charité Universitätsmedizin Berlin, Institute of Virology | Charité Universitätsmedizin Berlin, Institute of Virology | Barbara Mühlemann; Christian Drosten; Julia Schneider; Julia Tesch; Jörn Beheim-Schwarzbach; Talitha Veith; Terry Jones; Tobias Bleicker; Victor M Corman |
| EPI_ISL_7337463, EPI_ISL_7337464, EPI_ISL_7337465, EPI_ISL_7337466, EPI_ISL_7337468, EPI_ISL_7337469, EPI_ISL_7337470, EPI_ISL_7337471, EPI_ISL_7337472, EPI_ISL_7337473, EPI_ISL_7337474, EPI_ISL_7337475, EPI_ISL_7337476, EPI_ISL_7337477, EPI_ISL_7337478, EPI_ISL_7337479, EPI_ISL_7337480, EPI_ISL_7337481, EPI_ISL_7337482, EPI_ISL_7337483, EPI_ISL_7337484, EPI_ISL_7337485, EPI_ISL_7337486, EPI_ISL_7337487, EPI_ISL_7337488, EPI_ISL_7337489, EPI_ISL_7337490, EPI_ISL_7337495, EPI_ISL_7337496, EPI_ISL_7337497, EPI_ISL_7337498, EPI_ISL_7337499, EPI_ISL_7337500, EPI_ISL_7337501, EPI_ISL_7337502, EPI_ISL_7337503, EPI_ISL_7337504, EPI_ISL_7337505, EPI_ISL_7337506, EPI_ISL_7337507, EPI_ISL_7337508, EPI_ISL_7337509, EPI_ISL_7337510, EPI_ISL_7337511 | | | |
| see above | Charlotte Maxeke Johannesburg Academic Hospital | National Institute for Communicable Diseases of the National Health Laboratory Service | Amoako DG; Bhiman JN; Everatt J; Ismail A; Mahlangu B; Mnguni A; Mohale T; Ntuli N; Scheepers C |
| EPI_ISL_7337512, EPI_ISL_7337513, EPI_ISL_7337515, EPI_ISL_7337516, EPI_ISL_7337517, EPI_ISL_7337518, EPI_ISL_7337520, EPI_ISL_7337521, EPI_ISL_7337524, EPI_ISL_7337525, EPI_ISL_7337527, EPI_ISL_7337528, EPI_ISL_7337531, EPI_ISL_7337532, EPI_ISL_7337533, EPI_ISL_7337534, EPI_ISL_7337535, EPI_ISL_7337536, EPI_ISL_7337537, EPI_ISL_7337538, EPI_ISL_7337539, EPI_ISL_7337540, EPI_ISL_7337541, EPI_ISL_7337542, EPI_ISL_7337543, EPI_ISL_7337544, EPI_ISL_7337545, EPI_ISL_7337546, EPI_ISL_7337547, EPI_ISL_7337548, EPI_ISL_7337549, EPI_ISL_7337550, EPI_ISL_7337551, EPI_ISL_7337552, EPI_ISL_7337553, EPI_ISL_7337558, EPI_ISL_7337559, EPI_ISL_7337560, EPI_ISL_7337561, EPI_ISL_7337562, EPI_ISL_7337563, EPI_ISL_7337564 | | | |
| see above | Chris Hani Baragwanath Laboratory | National Institute for Communicable Diseases of the National Health Laboratory Service | Amoako DG; Bhiman JN; Everatt J; Ismail A; Mahlangu B; Mnguni A; Mohale T; Ntuli N; Scheepers C |
| EPI_ISL_7154390 | Clina-Lancet | National Reference Laboratory, Nigeria Centre for Disease Control | Catherine Okoi; Chimaobi Chukwu; Dr Ifedayo Adetifa; Dr Ndodo Nnaemeka; Dr Omoare Adesuyi; Nwando Mba; Olajumoke Babatunde; Olusola Anuoluwapo Akanbi; Oyeronke Ayansola |
| EPI_ISL_7265967, EPI_ISL_7265976, EPI_ISL_7265979, EPI_ISL_7266027, EPI_ISL_7266045, EPI_ISL_7266056, EPI_ISL_7266083, EPI_ISL_7266160 | | | |
| see above | Clinical Microbiology Laboratory, Tel Aviv Sourasky Medical Center | Clinical Microbiology Laboratory, Tel Aviv Sourasky Medical Center | Alon Ziv; Amos Adler; Katya Levytskyi; Lior Handler; Ora Halutz |
| EPI_ISL_7042161, EPI_ISL_7042168, EPI_ISL_7462220, EPI_ISL_7462233, EPI_ISL_7462310, EPI_ISL_7462311, EPI_ISL_7462312 | | | |
| see above | Clinical Virology | Clinical Bacteriology, University Hospital Basel | Adrian Egli; Alfredo Mari; Fanny Wegner; Hans Hirsch; Helena MB Seth-Smith; Julia Bielicki; Karoline Leuzinger; Manuel Battegay; Tim Roloff |
| EPI_ISL_7373598 | Clinical Virology, Children's Hospital Los Angeles | Clinical Virology, Children's Hospital Los Angeles | Alexander Judkins; Cheryl Pool; Javier Mestas; Jennifer Dien Bard; John Fissel; Maurice O'Gorman |
| EPI_ISL_7462438 | Cliniques universitaires Saint-Luc | UCLouvain/IREC/MBLG-CTMA | Benoit Kabamba Mukadi; Bertrand Bearzatto; Jean-Luc Gala; Nicolas Pinte; Paul Blanpain; Simon Ophélie; Valentin Coste |
| EPI_ISL_6951145 | Color Genomics | Chiu Laboratory, University of California, San Francisco | Alicia Sotomayor-Gonzalez; Alicia Zhou; Amy Garlin; Charles Chiu; Darpun Sachdev; Katherine Hernandez; Scott Topper; Susan Philip; Venice Servellita; Yueyuan Zhang |
| EPI_ISL_7010485 | Colorado Department of Public Health and Environment | Colorado Department of Public Health and Environment | Alexandria Rossheim; Diana Ir; Emily A. Travanty; Laura Bankers; Mandy Waters; Michael A. Martin; Molly C. Hetherington-Rauth; Sarah Elizabeth Totten; Shannon R. Matzinger |
| EPI_ISL_7451063, EPI_ISL_7451073, EPI_ISL_7451079, EPI_ISL_7451089 | Cruz Vermelha Portuguesa | Instituto Nacional de Saude (INSA) | Borges et al |
| EPI_ISL_7062087, EPI_ISL_7063588, EPI_ISL_7272249, EPI_ISL_7514034, EPI_ISL_7514991, EPI_ISL_7515043, EPI_ISL_7515374, EPI_ISL_7516043, EPI_ISL_7517471, EPI_ISL_7517646, EPI_ISL_7518103, EPI_ISL_7518795, EPI_ISL_7520214, EPI_ISL_7520543, EPI_ISL_7520819, EPI_ISL_7523657, EPI_ISL_7525859, EPI_ISL_7526218, EPI_ISL_7528707, EPI_ISL_7529040, EPI_ISL_7529817, EPI_ISL_7530609, EPI_ISL_7532336, EPI_ISL_7532391, EPI_ISL_7533005, EPI_ISL_7534407, EPI_ISL_7534541 | | | |
| see above | Department of Bacteria, Parasites and Fungi, Statens Serum Institut, Copenhagen, Denmark | Statens Serum Institut Bioinformatics and Microbial Genomics | Danish Covid-19 Genome Consortium |
| EPI_ISL_7042669 | Department of Clinical Microbiology | GIGA Medical Genomics | Bouchra Boujemla; Claire Gourzonès; Cécile Meex; Keith Durkin; Laurent Gillet; Maria Artesi; Marie-Pierre Hayette; Nadine Cambisano; Nathalie Renotte; Olivier Ek; Sébastien Bontems; Vincent Bours |
| EPI_ISL_7512899, EPI_ISL_7520649, EPI_ISL_7531709, EPI_ISL_7533537, EPI_ISL_7533749 | Department of Clinical Microbiology, Odense University Hospital, Odense, Denmark | Statens Serum Institut Bioinformatics and Microbial Genomics | Danish Covid-19 Genome Consortium |
| EPI_ISL_7192723, EPI_ISL_7192733, EPI_ISL_7192734, EPI_ISL_7485700 | Department of Health Technology and Informatics, The Hong Kong Polytechnic University | Department of Health Technology and Informatics, The Hong Kong Polytechnic University | Alan Ka-Lun Wu; Alex Yat-Man Ho; Barry Kin-Chung Wong; Chloe Toi-Mei Chan; David Ho-Keung Shum; Denise Sze-Hang Wong; Gilman Kit-Hang Siu; Hiu-Yin Lao; Hoi-Ching Jim; Ivan Tak-Fai Wong; Jake Siu-Lun Leung; Kam-Tong Yip; Kenneth Siu-Sing Leung; Kingsley King-Gee Tam; Kitty Sau-Chun Fung; Kristine Luk; Lam-Kwong Lee; Miranda Chong-Yee Yau; Sandy Ka-Yee Chau; Shea Ping Yip; Tak-Lun Que; Timothy Ting-Leung Ng; Wing Cheong Yam; Wing-Hei Lo; Wing-Kin To; Yvette Wai-Man Lai |
| EPI_ISL_6841980, EPI_ISL_6841981, EPI_ISL_7138045, EPI_ISL_7357684, EPI_ISL_7385702 | Department of Microbiology, The University of Hong Kong | Department of Microbiology, The University of Hong Kong | Kelvin K.W. To; Kwok-Yung Yuen |
| EPI_ISL_7201444 | Department of Virology and Immunology, University of Helsinki and Helsinki University Hospital, Huslab Finland | Department of Virology, Faculty of Medicine, University of Helsinki, Helsinki, Finland | Hanna Jarva; Hanna Liimatainen; Hanna Vauhkonen; Hussein Alburkat; Maija Lappalainen; Mert Erdin; Olli Vapalahti; Phuoc Truong; Ravi Kant; Sari Hannula; Satu Kurkela; Teemu Smura |
| EPI_ISL_6972689 | Dept. of Laboratory Medicine | Dept. of Laboratory Medicine | Claudia Weber; Fabian Konig; Harald Esterbauer; Oswald Wagner; Robert Strassl; Sabina Plumer; Victoria Six |

| | | | |
|---|---|---|---|
| EPI_ISL_7464539, EPI_ISL_7464543 | Dept. of Microbiology and Infection Control, Akershus University Hospital HF | Dept. of Microbiology and Infection Control, Akershus University Hospital HF | Alexander Hesselberg Løvestad; Hege Vangstein Aamot |
| EPI_ISL_7121080 | Diagnofirm Medical Laboratories | Botswana Harvard HIV Reference Laboratory | Boitumelo Zuze; Botshelo Radibe; Dorcas Maruapula; Joseph Makhema; Keoratile Ntshambiwa; Kgomotso Moruisi; Legodile Kooepile; Mosepele Mosepele; Mphaphi B. Mbulawa; Ontlametse T. Bareng; Pamela Smith-Lawrence; Roger Shapiro; Sefetogi Ramaologa; Shahin Lockman; Sikhulile Moyo; Simani Gaseitsiwe; Thongbotho Mphoyakgosi; Wonderful T. Choga |
| EPI_ISL_6959926, EPI_ISL_6959935, EPI_ISL_6959993, EPI_ISL_7406117, EPI_ISL_7406118, EPI_ISL_7406119, EPI_ISL_7406124, EPI_ISL_7406125, EPI_ISL_7406126 | | | |
| see above | Division of Emerging Infectious Diseases, Bureau of Infectious Diseases Diagnosis Control, Korea Disease Control and Prevention Agency | Division of Emerging Infectious Diseases, Bureau of Infectious Diseases Diagnosis Control, Korea Disease Control and Prevention Agency | Ae Kyung Park; Chae Young Lee; Eun-Jin Kim; Heui Man Kim; Hyuck Jin Lee; Il-Hwan Kim; Jeong-Ah Kim; Jeong-Min Kim |
| EPI_ISL_6842156, EPI_ISL_6842159, EPI_ISL_6842162, EPI_ISL_6842163, EPI_ISL_6842165, EPI_ISL_7452732, EPI_ISL_7452733, EPI_ISL_7452734, EPI_ISL_7452735, EPI_ISL_7452736, EPI_ISL_7452737, EPI_ISL_7452738, EPI_ISL_7452741, EPI_ISL_7452742, EPI_ISL_7452744, EPI_ISL_7452745, EPI_ISL_7452746, EPI_ISL_7452749, EPI_ISL_7452750, EPI_ISL_7452751, EPI_ISL_7452758, EPI_ISL_7452761, EPI_ISL_7452762, EPI_ISL_7452763, EPI_ISL_7452764, EPI_ISL_7452765, EPI_ISL_7452771, EPI_ISL_7452780, EPI_ISL_7452781, EPI_ISL_7452782, EPI_ISL_7452783, EPI_ISL_7452785, EPI_ISL_7452792, EPI_ISL_7452793, EPI_ISL_7452794, EPI_ISL_7452795, EPI_ISL_7452796, EPI_ISL_7452797, EPI_ISL_7452798, EPI_ISL_7452799, EPI_ISL_7452800, EPI_ISL_7456525, EPI_ISL_7456526, EPI_ISL_7544929 | | | |
| see above | Division of Medical Virology, National Health Laboratory Service (NHLS), Tygerberg Hospital / Stellenbosch University | Division of Medical Virology, National Health Laboratory Service (NHLS), Tygerberg Hospital / Stellenbosch University | Gert van Zyl; Kamela Mahlakwane; Shannon Wilson; Susan Engelbrecht; Tania Stander; Tongai Maponga; Wolfgang Preiser |
| EPI_ISL_7062590, EPI_ISL_7506397, EPI_ISL_7506422, EPI_ISL_7506490 | Dr. Risch Ostschweiz AG | Dr Risch Laboratory | Dominique Fabien Hilti; Faina Wehrli; Lorenz Risch; Martin Risch; Nadia Wohlwend; Sinem Kas; Thomas Bodmer |
| EPI_ISL_7469070 | Dutch COVID-19 response team | Medical Microbiology, Maastricht University Medical Centre | Brian van der Veer*; Carmen Reumkens; Christian Hoebe; Erik Beuken; Jozef Dingemans*; Lieke van Alphen; Paul Savelkoul |
| EPI_ISL_6841607, EPI_ISL_6841608, EPI_ISL_6841609, EPI_ISL_6841610, EPI_ISL_6841611, EPI_ISL_6841612, EPI_ISL_6841613, EPI_ISL_6841614, EPI_ISL_6841615, EPI_ISL_6841616, EPI_ISL_6841617, EPI_ISL_6841618, EPI_ISL_6841619, EPI_ISL_7470657, EPI_ISL_7471412, EPI_ISL_7471413, EPI_ISL_7471451, EPI_ISL_7471520, EPI_ISL_7471548, EPI_ISL_7471549, EPI_ISL_7471975 | | | |
| see above | Dutch COVID-19 response team | National Institute for Public Health and the Environment (RIVM) | Adam Meijer; AnneMarie van den Brandt; Annelies Kroneman; Bas van der Veer; Chantal Reusken; Dennis Schmitz; Dirk Eggink; Florian Zwagemaker; Harry Vennema; Ivo van Walle; Jeroen Cremer; Jil Kocken; Karim Hajji; Kim Freriks; Linda van Someren; Lisa Wijsman; Lynn Aarts; Ryanne Jaarsma; Sanne Bos; Sharon van den Brink; Stijn van Rossum; on behalf of the national COVID-19 response team |
| EPI_ISL_6826713, EPI_ISL_6826714, EPI_ISL_6989667, EPI_ISL_7015154, EPI_ISL_7160041, EPI_ISL_7160042 | Dynacare | National Microbiology Laboratory (NML) | Anna Majer; Anneliese Landgraff; CanCOGeN's metadata curation team; Darian Hole; Dynacare Brampton COVID-19 Diagnostic team; Elsie Grudeski; Gary Van Domselaar; Gordon Jolly; Grace Seo; Jennifer Tanner; Madison Chapel; Morag Graham; Natalie Knox; Nathalie Bastien; Philip Mabon; Public Health Agency of Canada CanCOGeN team; Rhiannon Huzarewich; Russell Mandes; Shari Tyson; Timothy Booth; Yan Li |
| EPI_ISL_6949582, EPI_ISL_7123666, EPI_ISL_7123680, EPI_ISL_7123692, EPI_ISL_7330020, EPI_ISL_7330021, EPI_ISL_7330161, EPI_ISL_7464335, EPI_ISL_7464340, EPI_ISL_7464346 | | | |
| see above | Edmonton Provincial Lab | Alberta Precision Labs (APL) | Buss; Croxen M; Deo A; Dieu P; E; Ferrato C; Gill K; Khan F; Koleva P; Li V; Lloyd C; Lynch T; Ma R; Murphy S; Pabbaraju K; Shokoples S; Thayer J; Tipples G; Whitehouse M; Wong A; Yu C; Zelyas N |
| EPI_ISL_7547732, EPI_ISL_7547733 | Everight diagnostics (Abuja) | National Reference Laboratory, Nigeria Centre for Disease Control | Catherine Okoi; Chimaobi Chukwu; Dr Ifedayo Adetifa; Dr Ndodo Nnaemeka; Dr Omoare Adesuyi; Nwando Mba; Olajumoke Babatunde; Olusola Anuoluwapo Akanbi; Oyeronke Ayansola |
| EPI_ISL_7313687 | Florida Bureau of Public Health Laboratories | Florida Bureau of Public Health Laboratories | Jason Blanton; Namratha Tarigopula; Sarah Schmedes; Tiffany Splatt |
| EPI_ISL_7154399, EPI_ISL_7497709, EPI_ISL_7497711, EPI_ISL_7497739, EPI_ISL_7497740, EPI_ISL_7497741, EPI_ISL_7497742, EPI_ISL_7497767 | | | |
| see above | Fulgent Genetics | Centers for Disease Control and Prevention Division of Viral Diseases, Pathogen Discovery | Becky Tsai; Benafsh Sapra; Benjamin Rambo-Martin; Christopher Gulvick; Clinton Paden; Dakota Howard; Dhwani Batra; Doreen Ng; Duncan MacCannell; Erisa Sula; Harry Gao; James Xie; Jason Caravas; John Gao; Joseph Fierro; Kristine Lacek; Matthew Schmerer; Mickey Li; Peter Cook; Scott Sammons; Shatavia Morrison; Tymeckia Kendall; Victoria Caban Figueroa; Yan Meng; Yvette Unoarumhi |
| EPI_ISL_6971572, EPI_ISL_6989155, EPI_ISL_7248772, EPI_ISL_7248778, EPI_ISL_7248797, EPI_ISL_7248805, EPI_ISL_7248814, EPI_ISL_7248821, EPI_ISL_7470261 | | | |
| see above | Furst Medical Laboratory | Norwegian Institute of Public Health, Department of Virology | Atiya R Ali; Debech Nadia; Engebretsen Serina Beate; Garcia Llorente Ignacio; Hilde Elshaug; Hilde Vollan; Jon Bråte; Kamilla Heddeland Instefjord; Karoline Bragstad; Kathrine Stene-Johansen; Line Victoria Moen; Marie Paulsen Madsen; Olav Hungnes; Pedersen Benedikte Nevjen; Rasmus Riis Kopperud |
| EPI_ISL_7473158 | GA Department of Public Health | GA Department of Public Health | Aliyah Fields; Jonathan Edwards; Sharmila Talekar; Stacy Reeves; Taylor Smith; Tonia Parrott |
| EPI_ISL_7173962 | GENEPATH | NCL, Pune | Ajinkya Khilari; Anu Raghunathan; Bhagyashree Litikar; Dhanasekaran Shanmugam; Divya Niveditha; Jugal Kanekar; Shikha Takur |
| EPI_ISL_7313494, EPI_ISL_7478195, EPI_ISL_7478220, EPI_ISL_7478233 | GH A.CHENEVIER-H.MONDOR | Department of Virology, Henri Mondor University Hospital, Assistance Publique Hôpitaux de Paris, Université Paris-Est Créteil, INSERM U955 | Alexandre Soulier; Christophe Rodriguez; Elisabeth Trawinski; Guillaume Gricourt; Jean-Michel Pawlotsky; Melissa N'Debi; Slim Fourati; Vanessa Demontant |
| EPI_ISL_6892613, EPI_ISL_6892620, EPI_ISL_6892631, EPI_ISL_6892639, EPI_ISL_6892644, EPI_ISL_6892650, EPI_ISL_6892653, EPI_ISL_6892659, EPI_ISL_6892661, EPI_ISL_6892665, EPI_ISL_6892674, EPI_ISL_6892683, EPI_ISL_6892693 | | | |
| see above | Germano de Sousa | Instituto Nacional de Saude (INSA) | Borges et al |
| EPI_ISL_7545349, EPI_ISL_7545360, EPI_ISL_7545585 | Gibraltar Health Authority Lab | Gibraltar Health Authority Covid-19 Laboratory | Bruna Martins; Dr Daniel Cassaglia; Dr Martyn Bell; Dr Nicholas Cortes; Dr Zoe Vincent; Sofia Lavelle |
| EPI_ISL_7549110, EPI_ISL_7549111 | Gravity Diagnostics, LLC | Gravity Diagnostics, LLC | Gravity Diagnostics |
| EPI_ISL_7544692 | Great Brak River Clinic wc GBC | NHLS/UCT | Arash Iranzadeh; Bruna Galvao; Carolyn Williamson; Deelan Doolabh; Diana Hardie; Gert Marais; Innocent Mudau; Luicer Olubayo; Lynn Tyers; Marvin Hsiao; Nokuzola Mbhele; Rageema Joseph; Stephen Korsman |
| EPI_ISL_7543874, EPI_ISL_7544059, EPI_ISL_7544158, EPI_ISL_7544262, EPI_ISL_7544652 | Groote Schuur Hospital wc GSH | NHLS/UCT | Arash Iranzadeh; Bruna Galvao; Carolyn Williamson; Deelan Doolabh; Diana Hardie; Gert Marais; Innocent Mudau; Luicer Olubayo; Lynn Tyers; Marvin Hsiao; Nokuzola Mbhele; Rageema Joseph; Stephen Korsman |
| EPI_ISL_6913917 | Grupo CR Diagnosticos | Instituto Adolfo Lutz Strategic Laboratory | Claudio Tavares Sacchi; Karoline Rodrigues Campos |
| EPI_ISL_6900139, EPI_ISL_6900141, EPI_ISL_6900142, EPI_ISL_6900143 | HELEN JOSEPH LABORATORY | National Institute for Communicable Diseases of the National Health Laboratory Service | Amoako DG; Bhiman JN; Everatt J; Ismail A; Mahlangu B; Mnguni A; Mohale T; Ntuli N; Scheepers C |
| EPI_ISL_6698790 | HOME QUARANTINE TASKFORCE | Hong Kong Department of Health | Alan K.L. Tsang; Edman T.K. Lam; Ken H.L. Ng; Peter C.W. Yip; Rickjason C.W. Chan |
| EPI_ISL_7045214 | HOSPITAL UNIVERSITARI DE BELLVITGE | Microbiology Department | Aida Gonzalez-Diaz; Anna Carrera-Salinas Yolanda Hernandez; Carmen Ardanuy; Jordi Camara; Jordi Niubó; Laura Calatayud; M Angeles Domínguez; Sara Marti |
| EPI_ISL_7470201 | Haukeland University Hospital, Dept. of Microbiology | Norwegian Institute of Public Health, Department of Virology | Atiya R Ali; Debech Nadia; Engebretsen Serina Beate; Garcia Llorente Ignacio; Hilde Elshaug; Hilde Vollan; Jon Bråte; Kamilla Heddeland Instefjord; Karoline Bragstad; Kathrine Stene-Johansen; Line Victoria Moen; Marie Paulsen Madsen; Olav Hungnes; Pedersen Benedikte Nevjen; Rasmus Riis Kopperud |
| EPI_ISL_7142185, EPI_ISL_7142260, EPI_ISL_7184579 | Health Services Laboratories | Wellcome Sanger Institute for the COVID-19 Genomics UK (COG-UK) Consortium | Cordelia Langford; David K. Jackson; Dominic Kwiatkowski; Ewan Harrison; Health Services Laboratories and Alex Alderton; Ian Johnston; Jeffrey Barrett; John Sillitoe on behalf of the Wellcome Sanger Institute COVID-19 Surveillance Team; Roberto Amato; Sonia Goncalves |
| EPI_ISL_7544466 | Helderberg Hospital wc HHH | NHLS/UCT | Arash Iranzadeh; Bruna Galvao; Carolyn Williamson; Deelan Doolabh; Diana Hardie; Gert Marais; Innocent Mudau; Luicer Olubayo; Lynn Tyers; Marvin Hsiao; Nokuzola Mbhele; Rageema Joseph; Stephen Korsman |
| EPI_ISL_7337440, EPI_ISL_7337441, EPI_ISL_7337442, EPI_ISL_7337443, EPI_ISL_7337444, EPI_ISL_7337445, EPI_ISL_7337446, EPI_ISL_7337447, EPI_ISL_7337448, EPI_ISL_7337449, EPI_ISL_7337450, EPI_ISL_7337451, EPI_ISL_7337452, EPI_ISL_7337453, EPI_ISL_7337454, EPI_ISL_7337455, EPI_ISL_7337458, EPI_ISL_7337460, EPI_ISL_7337461, EPI_ISL_7337462 | | | |
| see above | Helen Joseph Laboratory | National Institute for Communicable Diseases of the National Health Laboratory Service | Amoako DG; Bhiman JN; Everatt J; Ismail A; Mahlangu B; Mnguni A; Mohale T; Ntuli N; Scheepers C |
| EPI_ISL_7497723 | Helix | Centers for Disease Control and Prevention Division of Viral Diseases, Pathogen Discovery | Benjamin Rambo-Martin; Christopher Gulvick; Clinton Paden; Dakota Howard; Dhwani Batra; Duncan MacCannell; Erisa Sula; Helix CA; Jason Caravas; Kristine Lacek; Matthew Schmerer; Peter Cook; Scott Sammons; Shatavia Morrison; Tymeckia Kendall; Victoria Caban Figueroa; Yvette Unoarumhi |
| EPI_ISL_7265236, EPI_ISL_7265237, EPI_ISL_7462215 | Histopath | NSW Health Pathology - Institute of Clinical Pathology and Medical Research; Westmead Hospital; University of Sydney | Arnott A.; Draper J.; Gall M.; Martinez E.; Rockett R.; Sintchenko V.; on behalf of ICPMR |
| EPI_ISL_6590782, EPI_ISL_6832108 | Home Quarantine Taskforce | Hong Kong Department of Health | Alan K.L. Tsang; Edman T.K. Lam; Ken H.L. Ng; Peter C.W. Yip; Rickjason C.W. Chan |

| | | | |
|---|---|---|---|
| EPI_ISL_6716890, EPI_ISL_6716902 | Hong Kong Department of Health | School of Public Health, The University of Hong Kong | Dominic N.C. Tsang; Haogao Gu; Leo L.M. Poon; Malik Peiris |
| EPI_ISL_7373061, EPI_ISL_7373165, EPI_ISL_7373230 | Hospital General Universitario Albacete | Hospital General Universitario Albacete | Caridad Sainz de Baranda Camino; Lorena Robles-Fonseca |
| EPI_ISL_6851526, EPI_ISL_6902675, EPI_ISL_6971860, EPI_ISL_7042252 | Hospital General Universitario Gregorio Marañón | Hospital General Universitario Gregorio Marañón | Cristina Rodriguez-Grande; Darío García de Viedma; Jorge Rodríguez-Grande; Julia Suárez; Laura Pérez-Lago; Marta Herranz Martin; Patricia Muñoz; Pedro Sola Campoy; Pilar Catalán; Sergio Buenestado Serrano; Victor Manuel de la Cueva |
| EPI_ISL_7204336 | Hospital Universitari Dr. Josep Trueta | Institut d'Investigació Biomèdica de Girona \| Hospital Universitari Dr. Josep Trueta | Bernat del Olmo; Mel·lina Pinsach; Meritxell Deulofeu; Nuria Esther Neto; Paula Costa |
| EPI_ISL_7050911, EPI_ISL_7050918, EPI_ISL_7406515 | Hospital Universitari Vall d'Hebron - Vall d'Hebron Institut de Recerca | Hospital Universitari Vall d'Hebron - Vall d'Hebron Institut de Recerca | Alejandra González-Sánchez; Andrés Antón; Ariadna Rando; Carla Castillo; Cristina Andrés; Damir Garcia-Cehic; Josep Quer; Juliana Esperalba; Karen García; Maria Carmen Martin; Maria Gema Codina; Maria Piñana; Rodrigo Vásquez; Tomàs Pumarola |
| EPI_ISL_7277268 | Hospital Universitario Son Espases | Hospital Universitario Son Espases | Dr. Antonio Oliver; Dr. Carla López-Causapé; Dr. Gabriel Cabot; Hospital Universitario Son Espases; on behalf of Servicio de Microbiología |
| EPI_ISL_7329633 | Hospital Universitario de Guadalajara | Hospital General Universitario de Ciudad Real | Cristina Colmenarejo; José Martínez-Alarcón; Lidia García-Agudo; Marta Torres-Narbona; Soledad Illescas Fernández-Bermejo |
| EPI_ISL_7156454 | Hospital Ángeles Lomas | Instituto de diagnóstico y Referencia Epidemiologicos (INDRE) | Abril Rodriguez-Maldonado; Ariadna Medina-Benitez; Armando Rojo; Claudia Wong-Arambula; Ernesto Ramirez-Gonzalez; Fernando Gonzalez-Dominguez; Gisela Barrera-Badillo; Irma Lopez-Martinez; Joaquin Quiroz-Mercado; Leonardo Medina Arias; Lucia Hernandez-Rivas; Maribel Gonzalez-Villa; Natividad Cruz-Ortiz; Pilar Escamilla Llano; Raymundo Rodríguez Sandoval; Tatiana Nunez-Garcia; Vanessa Rivero-Arredondo |
| EPI_ISL_7415721, EPI_ISL_7415723, EPI_ISL_7415731, EPI_ISL_7415752, EPI_ISL_7415767, EPI_ISL_7415770, EPI_ISL_7415774, EPI_ISL_7415830, EPI_ISL_7415887 | | | |
| see above | Houston Methodist Hospital | Houston Methodist Hospital | Ilya J. Finkelstein; James J. Davis; Jessica Cambric; Jimmy Gollihar; Kristina Reppond; Layne Pruitt; Madison N. Shyer; Marcus Nguyen; Matthew Ojeda Saavedra; Paul A. Christensen; Prasanti Yerramilli; Randall J. Olsen; Robert Olson; Ryan Gadd; S. Wesley Long; Sishir Subedi; and James M. Musser |
| EPI_ISL_7210427 | Hrvatski zavod za javno zdravstvo | Hrvatski zavod za javno zdravstvo | Anita Jurić; Dragan Jurić; Irena Tabain; Ivana Ferenčak; Josipa Kuzle; Ljiljana Žmak; Mihaela Obrovac |
| EPI_ISL_7154340, EPI_ISL_7156753, EPI_ISL_7308635, EPI_ISL_7308771, EPI_ISL_7308875, EPI_ISL_7381064 | IHU Mediterranee Infection | IHU Mediterranee Infection | Philippe Colson et al. |
| EPI_ISL_7439547, EPI_ISL_7439558, EPI_ISL_7439622, EPI_ISL_7439646, EPI_ISL_7439650, EPI_ISL_7439657, EPI_ISL_7439722, EPI_ISL_7439730, EPI_ISL_7439754, EPI_ISL_7439781, EPI_ISL_7439806, EPI_ISL_7439827, EPI_ISL_7439832, EPI_ISL_7439902, EPI_ISL_7439935 | | | |
| see above | IMD - MVZ Labor Martinsried | Robert Koch Institute | |
| EPI_ISL_7451030, EPI_ISL_7451040, EPI_ISL_7451054, EPI_ISL_7451096 | INSA | Instituto Nacional de Saude (INSA) | Borges et al |
| EPI_ISL_7496734 | Idaho Bureau of Laboratories | Idaho Bureau of Laboratories | "R. Beukelman; Aimee Ceniseros; Christian Loera; Christopher Ball"; Matthew Charles Burns; Robert L. Voermans |
| EPI_ISL_7544430 | Ikhwezi CDC wc IKW | NHLS/UCT | Arash Iranzadeh; Bruna Galvao; Carolyn Williamson; Deelan Doolabh; Diana Hardie; Gert Marais; Innocent Mudau; Luicer Olubayo; Lynn Tyers; Marvin Hsiao; Nokuzola Mbhele; Rageema Joseph; Stephen Korsman |
| EPI_ISL_7381102 | Indian Council of Medical Research-National Institute of Virology, Microbial Containment Complex | Indian Council of Medical Research-National Institute of Virology, Microbial Containment Complex | Pragya D. Yadav |
| EPI_ISL_7166400 | Indira Gandhi Memorial Hospital | Indira Gandhi Memorial Hospital | D. Fathmath Nazla Rafeeq; Dr. Ibrahim Afzal; Mr. Ibrahim Nishan Ahmed; Ms. Aishath Shuhudha; Ms. Aminath Shazleena Abdul Rahman |
| EPI_ISL_7497749 | Infinity Biologix | Centers for Disease Control and Prevention Division of Viral Diseases, Pathogen Discovery | Benjamin Rambo-Martin; Chirayu Goswami; Christian Bixby; Christopher Gulvick; Clinton Paden; Dakota Howard; Dhwani Batra; Duncan MacCannell; Erisa Sula; Jason Caravas; Jonathan Schultz; Kristine Lacek; Matthew Schmerer; Peter Cook; Robin Grimwood; Russ Hager; Scott Sammons; Shatavia Morrison; Tymeckia Kendall; Victoria Caban Figueroa; Yihe Wang; Yvette Unoarumhi |
| EPI_ISL_7405329 | Inselspital Bern (Covid-Track) | Institute for Infectious Diseases, University of Bern | Alban Ramette; Christian Baumann; Cora Sägesser; Franziska Suter-Riniker; Loïc Borcard; Miguel A Terrazos Miani; Nicole Liechti; Pascal Bittel; Peter Keller; Sonja Gempeler; Stefan Neuenschwander; Stephen L Leib |
| EPI_ISL_7117396 | Institut für Labormedizin Mikrobiologie und Hygiene | Robert Koch Institute | |
| EPI_ISL_6832737 | Institute for Medical Virology Frankfurt | Institute for Medical Virology Frankfurt | Ciesek S.; Toptan T. |
| EPI_ISL_6959868, EPI_ISL_6959869, EPI_ISL_6959870, EPI_ISL_6959871, EPI_ISL_6959872, EPI_ISL_6959873, EPI_ISL_6959874, EPI_ISL_7479163, EPI_ISL_7479173, EPI_ISL_7479181, EPI_ISL_7479186, EPI_ISL_7479193, EPI_ISL_7479200, EPI_ISL_7479206 | | | |
| see above | Institute for Medical Virology, Frankfurt | Institute for Medical Virology, Frankfurt | Ciesek S.; Toptan T. |
| EPI_ISL_7404462, EPI_ISL_7404463 | Institute of Epidemiology, Disease Control and Research (IEDCR) | IEDCR-ideSHi Genomics Lab | Firdausi Qadri; Hassan Afrad; Manjur Hossain Khan; Omar Hamza; Tahmina Shirin |
| EPI_ISL_7507055 | Institute of Medical Science, University of Tokyo | Center for Influenza and Respiratory Virus Research, National Institute of Infectious Diseases (NIID) | Emi Takashita; Hideka Miura; Seiichiro Fujisaki; Yoshihiro Kawaoka; Yuko Sakai-Tagawa |
| EPI_ISL_6825546, EPI_ISL_6902052, EPI_ISL_6902053 | Institute of Virology, Department of Hygiene, Microbiology and Public Health at Innsbruck Medical University | Institute of Virology, Department of Hygiene, Microbiology and Public Health at Innsbruck Medical University | Andreas Aufschnaiter; Barbara Falkensammer; David Bante; Dorothee von Laer; Heribert Stoiber; Lukas Perro; Stephan Amstler; Wegene Borena |
| EPI_ISL_7550075 | Instituto Adolfo Lutz - Regional de Rio Claro | Instituto Adolfo Lutz, Interdiciplinary Procedures Center, Strategic Laboratory | Claudio Tavares Sacchi; Karoline Rodrigues Campos |
| EPI_ISL_7400617 | Iressef Genomics lab | IRESSEF | Abdou PADANE; Ambroise AHOUIDI; Aminata DIA; Aminata MBOUP; Astou Gaye GAYE; Barada CISSE; Birahim Piere NDIAYE; Cyrille Diedhiou; Diabou Diagne; Gora LO; Khadim GUEYE; Moustapha MBOW; Nafisatou LEYE; Ndeye Coumba Toure KANE; Papa Alassane DIAW; Samba Ndiour; Seni Ndiaye; Souleymane MBOUP; Yacine DIA |
| EPI_ISL_7160424 | Johns Hopkins Hospital Department of Pathology | Johns Hopkins Hospital Department of Pathology | Amary Fall; C. Paul Morris; David Gaston; Heba H. Mostafa; Julie M. Norton; Matthew Schwartz; Michael Forman; Raghda Eldesouki |
| EPI_ISL_6794907, EPI_ISL_6989250, EPI_ISL_7413964, EPI_ISL_7495278, EPI_ISL_7495279, EPI_ISL_7495280, EPI_ISL_7495281, EPI_ISL_7495282, EPI_ISL_7495283, EPI_ISL_7495284, EPI_ISL_7495285 | | | |
| see above | KU Leuven, Rega Institute, Clinical and Epidemiological Virology | KU Leuven, Rega Institute, Clinical and Epidemiological Virology | Bert Vanmechelen; Casper Geenen; Emmanuel André; Guy Baele; Joan Marti-Carerras; Joren; Lize Cuypers; Piet Maes; Raymenants; Sarah Gorissen; Simon Dellicour; Tony Wawina-Bokalanga |
| EPI_ISL_7219990, EPI_ISL_7220437, EPI_ISL_7220444, EPI_ISL_7286284 | Karolinska University Hospital Huddinge | Karolinska University Hospital Huddinge | Annika Tiveljung Lindell; Henning Onsbring; Jan Albert; Karina Hentrich; Lynda Eneh; Maria Ropat; Martin Ekman; Natalija Gerasimcik; Robert Dyrdak; Sandra Broddesson; Shambhu Ganeshappa Aralaguppe; Tanja Normark; Tobias Allander; Valtteri Wirta; Zhibing Yun |
| EPI_ISL_7381109, EPI_ISL_7381114, EPI_ISL_7457425, EPI_ISL_7457426 | Karolinska University Hospital Solna | Karolinska University Hospital Huddinge | Annika Tiveljung Lindell; Henning Onsbring; Jan Albert; Karina Hentrich; Lynda Eneh; Maria Ropat; Martin Ekman; Natalija Gerasimcik; Robert Dyrdak; Sandra Broddesson; Shambhu Ganeshappa Aralaguppe; Tanja Normark; Tobias Allander; Valtteri Wirta; Zhibing Yun |
| EPI_ISL_7433816, EPI_ISL_7443687, EPI_ISL_7443713 | Klinikum Ernst von Bergmann gemeinnützige GmbH - stationärer Bereich | Robert Koch Institute | |
| EPI_ISL_7224971 | LACEN do Distrito Federal | Instituto Adolfo Lutz Strategic Laboratory | Claudio Tavares Sacchi; Karoline Rodrigues Campos; Marlon Benedito Nascimento Santos |
| EPI_ISL_7550076 | LACEN do Distrito Federal | Instituto Adolfo Lutz, Interdiciplinary Procedures Center, Strategic Laboratory | Claudio Tavares Sacchi; Karoline Rodrigues Campos |
| EPI_ISL_6647956, EPI_ISL_6647957, EPI_ISL_6647958, EPI_ISL_6647959, EPI_ISL_6647960, EPI_ISL_6647961, EPI_ISL_6647962, EPI_ISL_6698792, EPI_ISL_6704863, EPI_ISL_6704864, EPI_ISL_6704865, EPI_ISL_6704866, EPI_ISL_6704867, EPI_ISL_6704868, EPI_ISL_6704869, EPI_ISL_6704870, EPI_ISL_6704871, EPI_ISL_6704872, EPI_ISL_6704873, EPI_ISL_6704874, EPI_ISL_6704875, EPI_ISL_6704876 | | | |
| see above | LANCET LABORATORY | National Institute for Communicable Diseases of the National Health | Amoako DG; Bhiman JN; Everatt J; Glass A; Ismail A; Mahlangu B; Mnguni A; Mohale T; Ntuli N; Scheepers C; Viana R; Wolter N |

| | | | |
|---|---|---|---|
| | | Laboratory Service | |
| EPI_ISL_6901960, EPI_ISL_6901961, EPI_ISL_7473154 | LATE - Laboratório de Técnicas Especiais - Hospital Israelita Albert Einstein | LATE - Laboratório de Técnicas Especiais - Hospital Israelita Albert Einstein | Alexandre Hideaki Takara; Ana Paula Moreira Salles; Anelisie da Silva Santos; Deyvid Amgarten; Erick Gustavo Dorlass; Fernanda de Mello Malta; João Renato Rebello Pinho; Luiz Vicente Rizzo; Marcio Anunciacao Menezes; Pedro Henrique Sebe Rodrigues; Raquel Riyuzo |
| EPI_ISL_7472277 | LKO | Jessa | Severine Berden et al. on behalf of the Jessa_cmdLab |
| EPI_ISL_7456529 | LSUHS Emerging Viral Threat Lab | LSUHS Emerging Viral Threat Laboratory | Adrian Almodovar; Alexander Mijalis; Andrew D. Yurochko; Christopher G. Kevil; Gregory L. Ware; Jennifer L. Carroll; Jeremy P. Kamil; John A. Vanchiere; Krista Queen; Maarten Van Diest; Rona S. Scott |
| EPI_ISL_7339434, EPI_ISL_7339435, EPI_ISL_7339436 | Lab voor klinische biologie | Lab voor klinische biologie | Bruno Verhasselt; Hannelore Hamerlinck; Marija Janevska; May-Linh Truong |
| EPI_ISL_7437787 | Labor Prof. Dr. G. Enders MVZ GbR | Robert Koch Institute | |
| EPI_ISL_7450035, EPI_ISL_7450728 | Laboratoire BIORANCE | CHU Pontchaillou | DE TAYRAC Marie; DENOUAL Florent; ETCHEVERRY Amandine; FEBREAU Christine; GALIBERT Marie Dominique; GROLHIER Claire; JAGLINE Steven; PRONIER Charlotte; QUENET Benjamin; SASSI Mohamed; THIBAULT Vincent |
| EPI_ISL_6971091 | Laboratoire analyse med | LABORIZON CENTRE BIOGROUP | HAGUENOER EVE; HOLSTEIN ANNE; JIMENEZ MELANIE; LEFLEUTER NICOLAS; POITIRON GREGOIRE |
| EPI_ISL_7309168 | Laboratorio Central de Saude Publica do Rio Grande do Sul/Centro Estadual de Vigilancia em Saude | Centro de Desenvolvimento Cientifico e Tecnologico (CDCT)/Centro Estadual de Vigilancia em Saude | Alana Rossetti; Cláudia Maria Dornelles da Silva; Fernanda Marques Godinho; Larissa Vitoria da Silva; Ludmila Fiorenzano Baethgen; Miguel S Andrade.; Regina Bones Barcellos; Richard Steiner Salvato; Tatiana Schaffer Gregianini |
| EPI_ISL_7497691 | Laboratory Corporation of America | Centers for Disease Control and Prevention Division of Viral Diseases, Pathogen Discovery | Amanda Douglas; Amanda Suchanek; Andrea Throop; Ayla Burns; Benjamin Rambo-Martin; Bobbi Croy; Brian Krueger; Brian Norvell; Christopher Gulvick; Christos Petropoulos; Clinton Paden; Craig Lukasik; Dakota Howard; Debbie Boles; Dhwani Batra; Duncan MacCannell; Eyad Almasri; Goran Stevovic; Howard Engler; Hrushikesh Deshmukh; Jake Humphrey; Jana Schroth; Jason Caravas; Joe Voshell; John Pruitt; Jonathan Meltzer; Jonathan Williams; Kimberly Wagner; Kristine Lacek; Lax Iyer; Lisa Pfefferle; Lyndon Tilson; Manoj Jain; Marcia Eisenberg; Mary Cristobal; Mary Williamson; Matthew Robinson; Matthew Schmerer; Michael Levandoski; Mike Sapeta; Mindy Nye; Minoo Agarwal; Mohan Kolli; Nuthawin Charoensri; Oren Cohen; Peter Cook; Prashant Gupta; Qian Zeng; Rama Ghatti; Scott Parker; Scott Ryan; Scott Sammons; Shatavia Morrison; Stanley Letovsky; Steven Ragan; Suresh Selvaraju; Susan Countryman; Susan Hicks; Suzanne Dale; Thomas Urban; Tim Kuphal; Tricia Zwiefelhofer; Tymeckia Kendall; Victoria Caban Figueroa; Vincent Drouillon; Yvette Unoarumhi |
| EPI_ISL_7220176 | Laboratory of Clinical Virology Heraklion Crete | Laboratory of Clinical Virology Heraklion Crete | Alexandros Zafiropoulos; George Sourvinos |
| EPI_ISL_7543999 | Lady Michaelis CDC wc LMC | NHLS/UCT | Arash Iranzadeh; Bruna Galvao; Carolyn Williamson; Deelan Doolabh; Diana Hardie; Gert Marais; Innocent Mudau; Luicer Olubayo; Lynn Tyers; Marvin Hsiao; Nokuzola Mbhele; Rageema Joseph; Stephen Korsman |
| EPI_ISL_7264143 | Lahey Hospital | New England Biolabs | Abel, G.; B.W.; C.J.; Elfahal, M.; Flynn; Heim, K.; Karolides, M.; L. and Langhorst; Leger, P.; Michaels, L.; Pinet, K.; Skelton, T.; Sun |
| EPI_ISL_7543847, EPI_ISL_7543913, EPI_ISL_7543935, EPI_ISL_7543949, EPI_ISL_7543960, EPI_ISL_7544508, EPI_ISL_7544601, EPI_ISL_7544676, EPI_ISL_7544705 | | | |
| see above | Lancet | NHLS/UCT | Arash Iranzadeh; Bruna Galvao; Carolyn Williamson; Deelan Doolabh; Diana Hardie; Gert Marais; Innocent Mudau; Luicer Olubayo; Lynn Tyers; Marvin Hsiao; Nokuzola Mbhele; Rageema Joseph; Stephen Korsman |
| EPI_ISL_6913991, EPI_ISL_6913992, EPI_ISL_6913993, EPI_ISL_6913994, EPI_ISL_6913995, EPI_ISL_6913996, EPI_ISL_6913997, EPI_ISL_6913998, EPI_ISL_6913999, EPI_ISL_6914000, EPI_ISL_6914001, EPI_ISL_6914002, EPI_ISL_6914003, EPI_ISL_6914004, EPI_ISL_6914005, EPI_ISL_6914006, EPI_ISL_6914007 | | | |
| see above | Lancet Laboratories | National Institute for Communicable Diseases of the National Health Laboratory Service | Amoako DG; Bhiman JN; Everatt J; Ismail A; Mahlangu B; Mnguni A; Mohale T; Ntuli N; Scheepers C; Wolter N |
| EPI_ISL_7420297, EPI_ISL_7420298, EPI_ISL_7420371, EPI_ISL_7420381 | Landesgesundheitsamt Baden-Württemberg | Robert Koch Institute | |
| EPI_ISL_7543863 | Langa Clinic wc LAN | NHLS/UCT | Arash Iranzadeh; Bruna Galvao; Carolyn Williamson; Deelan Doolabh; Diana Hardie; Gert Marais; Innocent Mudau; Luicer Olubayo; Lynn Tyers; Marvin Hsiao; Nokuzola Mbhele; Rageema Joseph; Stephen Korsman |
| EPI_ISL_7348417, EPI_ISL_7348427 | Lifebrain Covid Labor GmbH | Lifebrain Covid Labor GmbH | Filip Sima |
| EPI_ISL_6821008, EPI_ISL_6916148, EPI_ISL_7023724, EPI_ISL_7144808, EPI_ISL_7144986, EPI_ISL_7148837, EPI_ISL_7187307, EPI_ISL_7200868, EPI_ISL_7290346, EPI_ISL_7293790, EPI_ISL_7293812, EPI_ISL_7293816, EPI_ISL_7293841, EPI_ISL_7293869, EPI_ISL_7293903, EPI_ISL_7294032, EPI_ISL_7294185, EPI_ISL_7296873, EPI_ISL_7301592, EPI_ISL_7301667, EPI_ISL_7305287, EPI_ISL_7344013, EPI_ISL_7344598, EPI_ISL_7344677, EPI_ISL_7346484, EPI_ISL_7351048, EPI_ISL_7351423, EPI_ISL_7351936, EPI_ISL_7353170, EPI_ISL_7382609, EPI_ISL_7383321, EPI_ISL_7385376, EPI_ISL_7389680, EPI_ISL_7392768, EPI_ISL_7397825, EPI_ISL_7484328, EPI_ISL_7488052, EPI_ISL_7490160, EPI_ISL_7491433, EPI_ISL_7491455, EPI_ISL_7491475, EPI_ISL_7492192, EPI_ISL_7492231, EPI_ISL_7511780, EPI_ISL_7514364, EPI_ISL_7514804, EPI_ISL_7515071, EPI_ISL_7515102, EPI_ISL_7515212, EPI_ISL_7515213, EPI_ISL_7515290, EPI_ISL_7515719, EPI_ISL_7516761, EPI_ISL_7535745, EPI_ISL_7535814, EPI_ISL_7535856, EPI_ISL_7535938, EPI_ISL_7536494, EPI_ISL_7536603, EPI_ISL_7537524, EPI_ISL_7538327, EPI_ISL_7539165, EPI_ISL_7539548, EPI_ISL_7539590, EPI_ISL_7539720, EPI_ISL_7541054 | | | |
| see above | Lighthouse Lab in Alderley Park | Wellcome Sanger Institute for the COVID-19 Genomics UK (COG-UK) Consortium | Cordelia Langford; David K. Jackson; Dominic Kwiatkowski; Ewan Harrison; Ian Johnston; Jacquelyn Wynn; Jeffrey Barrett; John Sillitoe on behalf of the Wellcome Sanger Institute COVID-19 Surveillance Team; Mairead Hyland; Roberto Amato; Sonia Goncalves; The Lighthouse Lab in Alderley Park and Alex Alderton |
| EPI_ISL_6869226, EPI_ISL_6869363, EPI_ISL_6869407, EPI_ISL_6869556, EPI_ISL_6873612, EPI_ISL_6920174, EPI_ISL_6926451, EPI_ISL_6926743, EPI_ISL_6927893, EPI_ISL_7019951, EPI_ISL_7022011, EPI_ISL_7027157, EPI_ISL_7027809, EPI_ISL_7029582, EPI_ISL_7031141, EPI_ISL_7031412, EPI_ISL_7035922, EPI_ISL_7036426, EPI_ISL_7145006, EPI_ISL_7145516, EPI_ISL_7148141, EPI_ISL_7148719, EPI_ISL_7158402, EPI_ISL_7158414, EPI_ISL_7158423, EPI_ISL_7158488, EPI_ISL_7158498, EPI_ISL_7158548, EPI_ISL_7292889, EPI_ISL_7292960, EPI_ISL_7294550, EPI_ISL_7294585, EPI_ISL_7295180, EPI_ISL_7295212, EPI_ISL_7295689, EPI_ISL_7296032, EPI_ISL_7296139, EPI_ISL_7296254, EPI_ISL_7296303, EPI_ISL_7297042, EPI_ISL_7297075, EPI_ISL_7297120, EPI_ISL_7299420, EPI_ISL_7299535, EPI_ISL_7299536, EPI_ISL_7299709, EPI_ISL_7299880, EPI_ISL_7302122, EPI_ISL_7302177, EPI_ISL_7302222, EPI_ISL_7302501, EPI_ISL_7302554, EPI_ISL_7303186, EPI_ISL_7303590, EPI_ISL_7303781, EPI_ISL_7304114, EPI_ISL_7304964, EPI_ISL_7305445, EPI_ISL_7305675, EPI_ISL_7305684, EPI_ISL_7305734, EPI_ISL_7305755, EPI_ISL_7305800, EPI_ISL_7305876, EPI_ISL_7305903, EPI_ISL_7305934, EPI_ISL_7306117, EPI_ISL_7306238, EPI_ISL_7306338, EPI_ISL_7306343, EPI_ISL_7306399, EPI_ISL_7343337, EPI_ISL_7343938, EPI_ISL_7343985, EPI_ISL_7344167, EPI_ISL_7344174, EPI_ISL_7345819, EPI_ISL_7345863, EPI_ISL_7345932, EPI_ISL_7345966, EPI_ISL_7345975, EPI_ISL_7346136, EPI_ISL_7346249, EPI_ISL_7346862, EPI_ISL_7346896, EPI_ISL_7347059, EPI_ISL_7347482, EPI_ISL_7350891, EPI_ISL_7351023, EPI_ISL_7351093, EPI_ISL_7352907, EPI_ISL_7353351, EPI_ISL_7355546, EPI_ISL_7355957, EPI_ISL_7355992, EPI_ISL_7356061, EPI_ISL_7356171, EPI_ISL_7356256, EPI_ISL_7384310, EPI_ISL_7389585, EPI_ISL_7392688, EPI_ISL_7392706, EPI_ISL_7394189, EPI_ISL_7394919, EPI_ISL_7397103, EPI_ISL_7397308, EPI_ISL_7397319, EPI_ISL_7397480, EPI_ISL_7510398, EPI_ISL_7510491, EPI_ISL_7510550, EPI_ISL_7510572, EPI_ISL_7510800, EPI_ISL_7510886, EPI_ISL_7510922, EPI_ISL_7510928, EPI_ISL_7510978, EPI_ISL_7511143, EPI_ISL_7511154, EPI_ISL_7511233, EPI_ISL_7511305, EPI_ISL_7511455, EPI_ISL_7511587, EPI_ISL_7511662, EPI_ISL_7511677, EPI_ISL_7511702, EPI_ISL_7511820, EPI_ISL_7511832, EPI_ISL_7511896, EPI_ISL_7511986, EPI_ISL_7517098, EPI_ISL_7517101, EPI_ISL_7517216, EPI_ISL_7518683, EPI_ISL_7519628, EPI_ISL_7520531, EPI_ISL_7521048, EPI_ISL_7522619, EPI_ISL_7522673, EPI_ISL_7531008, EPI_ISL_7531228, EPI_ISL_7531322, EPI_ISL_7532295, EPI_ISL_7535753, EPI_ISL_7536021, EPI_ISL_7536166, EPI_ISL_7536246, EPI_ISL_7536342, EPI_ISL_7536453, EPI_ISL_7536705, EPI_ISL_7536927 | | | |
| see above | Lighthouse Lab in Glasgow | Wellcome Sanger Institute for the COVID-19 Genomics UK (COG-UK) Consortium | Anna Dominiczak and Alex Alderton; Carol Clugston; Cordelia Langford; David Gray; David K. Jackson; Dominic Kwiatkowski; Ewan Harrison; Harper VanSteenhouse; Ian Johnston; Jeffrey Barrett; John Sillitoe on behalf of the Wellcome Sanger Institute COVID-19 Surveillance Team; Roberto Amato; Sonia Goncalves; Yumi Kasai |
| EPI_ISL_6818284, EPI_ISL_6833884, EPI_ISL_6834467, EPI_ISL_6915964, EPI_ISL_6919027, EPI_ISL_6920616, EPI_ISL_6923186, EPI_ISL_7022755, EPI_ISL_7023187, EPI_ISL_7023265, EPI_ISL_7023667, EPI_ISL_7023675, EPI_ISL_7023683, EPI_ISL_7023726, EPI_ISL_7023789, EPI_ISL_7023867, EPI_ISL_7024334, EPI_ISL_7024438, EPI_ISL_7024495, EPI_ISL_7024659, EPI_ISL_7024687, EPI_ISL_7024839, EPI_ISL_7024991, EPI_ISL_7025373, EPI_ISL_7026085, EPI_ISL_7026396, EPI_ISL_7026536, EPI_ISL_7026616, EPI_ISL_7026695, EPI_ISL_7026716, EPI_ISL_7026777, EPI_ISL_7026952, EPI_ISL_7028268, EPI_ISL_7028487, EPI_ISL_7029353, EPI_ISL_7033208, EPI_ISL_7037264, EPI_ISL_7143464, EPI_ISL_7146338, EPI_ISL_7146517, EPI_ISL_7146524, EPI_ISL_7146933, EPI_ISL_7147199, EPI_ISL_7157375, EPI_ISL_7157554, EPI_ISL_7181721, EPI_ISL_7182128, EPI_ISL_7182133, EPI_ISL_7182156, EPI_ISL_7185699, EPI_ISL_7185720, EPI_ISL_7185763, EPI_ISL_7185784, EPI_ISL_7185838, EPI_ISL_7185864, EPI_ISL_7185866, EPI_ISL_7187248, EPI_ISL_7187503, EPI_ISL_7188445, EPI_ISL_7188816, EPI_ISL_7188897, EPI_ISL_7188957, EPI_ISL_7189046, EPI_ISL_7189048, EPI_ISL_7189067, EPI_ISL_7189073, EPI_ISL_7196773, EPI_ISL_7197261, EPI_ISL_7197402, EPI_ISL_7197454, EPI_ISL_7198739, EPI_ISL_7198774, EPI_ISL_7198902, EPI_ISL_7199159, EPI_ISL_7202939, EPI_ISL_7202951, EPI_ISL_7202968, EPI_ISL_7202977, EPI_ISL_7202998, EPI_ISL_7203007, EPI_ISL_7203016, EPI_ISL_7203017, EPI_ISL_7203032, EPI_ISL_7203085, EPI_ISL_7203096, EPI_ISL_7203100, EPI_ISL_7203105, EPI_ISL_7203132, EPI_ISL_7203159, EPI_ISL_7203161, EPI_ISL_7203182, EPI_ISL_7203185, EPI_ISL_7203202, EPI_ISL_7203249, EPI_ISL_7203710, EPI_ISL_7203762, EPI_ISL_7203765, EPI_ISL_7203936, EPI_ISL_7204224, EPI_ISL_7293149, EPI_ISL_7293182, EPI_ISL_7293573, EPI_ISL_7293699, EPI_ISL_7293701, EPI_ISL_7293733, EPI_ISL_7293785, EPI_ISL_7296211, EPI_ISL_7297545, EPI_ISL_7297579, EPI_ISL_7300159, EPI_ISL_7300279, EPI_ISL_7300286, EPI_ISL_7300537, EPI_ISL_7300658, EPI_ISL_7300878, EPI_ISL_7300889, EPI_ISL_7300915, EPI_ISL_7300986, EPI_ISL_7301005, EPI_ISL_7301013, EPI_ISL_7301015, EPI_ISL_7301165, EPI_ISL_7301209, EPI_ISL_7301266, EPI_ISL_7301307, EPI_ISL_7301521, EPI_ISL_7301561, EPI_ISL_7301876, EPI_ISL_7304476, EPI_ISL_7304564, EPI_ISL_7304690, EPI_ISL_7304994, EPI_ISL_7305027, EPI_ISL_7305176, EPI_ISL_7340046, EPI_ISL_7340118, EPI_ISL_7340964, EPI_ISL_7341048, EPI_ISL_7344931, EPI_ISL_7345012, EPI_ISL_7345138, EPI_ISL_7345164, EPI_ISL_7346326, EPI_ISL_7346393, EPI_ISL_7346410, EPI_ISL_7346479, EPI_ISL_7346521, EPI_ISL_7346666, EPI_ISL_7346704, EPI_ISL_7346758, EPI_ISL_7346809, EPI_ISL_7346816, EPI_ISL_7346835, EPI_ISL_7347737, EPI_ISL_7347952, EPI_ISL_7347992, EPI_ISL_7349648, EPI_ISL_7349666, EPI_ISL_7349764, EPI_ISL_7349797, EPI_ISL_7349804, EPI_ISL_7350042, EPI_ISL_7350201, EPI_ISL_7350318, EPI_ISL_7350332, EPI_ISL_7350365, EPI_ISL_7350520, EPI_ISL_7350526, EPI_ISL_7350556, EPI_ISL_7350583, EPI_ISL_7353443, EPI_ISL_7353487, EPI_ISL_7353795, EPI_ISL_7353847, EPI_ISL_7353853, EPI_ISL_7353856, EPI_ISL_7353981, EPI_ISL_7354003, EPI_ISL_7354110, EPI_ISL_7354518, EPI_ISL_7354594, EPI_ISL_7354762, EPI_ISL_7354795, EPI_ISL_7354941, EPI_ISL_7354957, EPI_ISL_7354991, EPI_ISL_7355006, EPI_ISL_7355123, EPI_ISL_7381834, EPI_ISL_7381856, EPI_ISL_7381881, EPI_ISL_7382005, EPI_ISL_7382097, EPI_ISL_7382103, EPI_ISL_7382156, EPI_ISL_7382200, EPI_ISL_7382215, EPI_ISL_7382254, EPI_ISL_7382416, EPI_ISL_7383376, EPI_ISL_7383676, EPI_ISL_7383683, EPI_ISL_7383685, EPI_ISL_7383740, EPI_ISL_7383773, EPI_ISL_7383781, EPI_ISL_7383783, EPI_ISL_7383798, EPI_ISL_7383852, EPI_ISL_7383856, EPI_ISL_7383863, EPI_ISL_7383885, EPI_ISL_7383921, EPI_ISL_7383922, EPI_ISL_7383936, EPI_ISL_7383941, EPI_ISL_7383945, EPI_ISL_7383954, EPI_ISL_7383977, EPI_ISL_7383978, EPI_ISL_7383980, EPI_ISL_7384190, EPI_ISL_7384545, EPI_ISL_7384619, EPI_ISL_7385445, EPI_ISL_7385606, EPI_ISL_7386026, EPI_ISL_7386052, EPI_ISL_7386325, EPI_ISL_7386397, EPI_ISL_7390306, EPI_ISL_7390618, EPI_ISL_7391659, EPI_ISL_7391668, EPI_ISL_7391823, EPI_ISL_7393627, EPI_ISL_7393737, EPI_ISL_7393800, EPI_ISL_7393853, EPI_ISL_7393873, EPI_ISL_7395502, EPI_ISL_7395791, EPI_ISL_7395797, EPI_ISL_7395849, EPI_ISL_7395905, EPI_ISL_7396016, EPI_ISL_7396038, EPI_ISL_7396042, EPI_ISL_7396056, EPI_ISL_7396060, EPI_ISL_7396114, EPI_ISL_7396120, EPI_ISL_7396167, EPI_ISL_7396207, EPI_ISL_7396238, EPI_ISL_7396262, EPI_ISL_7396271, EPI_ISL_7396409, EPI_ISL_7396415, EPI_ISL_7396446, EPI_ISL_7396483, EPI_ISL_7396525, EPI_ISL_7396533, EPI_ISL_7396579, EPI_ISL_7396628, EPI_ISL_7396640, EPI_ISL_7396987, EPI_ISL_7398089, EPI_ISL_7398228, EPI_ISL_7398295, EPI_ISL_7398393, EPI_ISL_7478708, EPI_ISL_7478734, EPI_ISL_7478799, EPI_ISL_7478827, EPI_ISL_7479102, EPI_ISL_7479225, EPI_ISL_7479311, EPI_ISL_7479350, EPI_ISL_7479729, EPI_ISL_7480249, EPI_ISL_7480340, EPI_ISL_7480459, EPI_ISL_7480563, EPI_ISL_7480779, EPI_ISL_7480983, EPI_ISL_7481103, EPI_ISL_7481888, EPI_ISL_7483648, EPI_ISL_7483732, EPI_ISL_7483778, EPI_ISL_7483853, EPI_ISL_7484047, EPI_ISL_7484394, EPI_ISL_7484568, EPI_ISL_7484575, EPI_ISL_7484580, EPI_ISL_7484583, EPI_ISL_7484584, EPI_ISL_7484632, EPI_ISL_7484936, EPI_ISL_7485137, EPI_ISL_7485159, EPI_ISL_7485300, EPI_ISL_7485391, EPI_ISL_7485734, EPI_ISL_7485763, EPI_ISL_7485880, EPI_ISL_7485890, EPI_ISL_7485899, EPI_ISL_7485918, EPI_ISL_7486077, EPI_ISL_7486120, EPI_ISL_7486163, EPI_ISL_7486373, EPI_ISL_7486447, EPI_ISL_7486481, EPI_ISL_7486517, EPI_ISL_7486635, EPI_ISL_7486819, EPI_ISL_7486971, EPI_ISL_7487426, EPI_ISL_7487488, EPI_ISL_7487790, EPI_ISL_7487954, EPI_ISL_7488141, EPI_ISL_7488200, EPI_ISL_7488279, EPI_ISL_7489187, EPI_ISL_7489326, EPI_ISL_7489389, EPI_ISL_7489446, EPI_ISL_7489786, EPI_ISL_7489819, EPI_ISL_7489953, EPI_ISL_7489980, EPI_ISL_7490259, EPI_ISL_7490487, EPI_ISL_7490584, EPI_ISL_7490883, EPI_ISL_7490957, EPI_ISL_7509888, EPI_ISL_7509901, EPI_ISL_7509939, EPI_ISL_7510081, EPI_ISL_7510216, EPI_ISL_7510241, EPI_ISL_7511995, EPI_ISL_7512017, EPI_ISL_7512047, EPI_ISL_7512058, EPI_ISL_7512077, EPI_ISL_7512119, EPI_ISL_7512155, EPI_ISL_7512175, EPI_ISL_7512265, EPI_ISL_7512279, EPI_ISL_7512284, EPI_ISL_7512302, EPI_ISL_7512321, EPI_ISL_7512475, EPI_ISL_7512549, EPI_ISL_7512707, EPI_ISL_7512966, EPI_ISL_7513072, EPI_ISL_7513131, EPI_ISL_7513234, EPI_ISL_7514854, EPI_ISL_7515519, EPI_ISL_7515566, EPI_ISL_7515634, EPI_ISL_7515973, EPI_ISL_7516148, EPI_ISL_7517004, EPI_ISL_7519115, EPI_ISL_7520174, EPI_ISL_7520334, EPI_ISL_7520362, EPI_ISL_7520998, EPI_ISL_7522351, EPI_ISL_7522461, EPI_ISL_7523110, EPI_ISL_7523203, EPI_ISL_7523364, EPI_ISL_7523595, EPI_ISL_7524106, EPI_ISL_7524864, EPI_ISL_7525422, EPI_ISL_7526214, EPI_ISL_7526231, EPI_ISL_7526276, EPI_ISL_7526372, EPI_ISL_7526379, EPI_ISL_7526426, EPI_ISL_7526472, EPI_ISL_7526476, EPI_ISL_7526571, EPI_ISL_7526583, EPI_ISL_7526592, EPI_ISL_7526597, EPI_ISL_7526604, EPI_ISL_7526609, EPI_ISL_7526665, EPI_ISL_7526672, EPI_ISL_7526690, EPI_ISL_7526766, EPI_ISL_7526796, EPI_ISL_7526806, EPI_ISL_7526823, EPI_ISL_7526841, EPI_ISL_7526878, EPI_ISL_7526961, EPI_ISL_7526985, EPI_ISL_7526997, EPI_ISL_7527046, EPI_ISL_7527101, EPI_ISL_7527267, EPI_ISL_7527283, EPI_ISL_7527298, EPI_ISL_7527348, EPI_ISL_7527381, EPI_ISL_7527452, EPI_ISL_7528184, EPI_ISL_7528477, EPI_ISL_7528569, EPI_ISL_7528699, EPI_ISL_7528999, EPI_ISL_7529344, EPI_ISL_7529479, EPI_ISL_7529568, EPI_ISL_7529705, EPI_ISL_7529714, EPI_ISL_7529749, EPI_ISL_7529754, EPI_ISL_7530006, EPI_ISL_7530158, EPI_ISL_7530274, EPI_ISL_7530290, EPI_ISL_7530610, EPI_ISL_7532349, EPI_ISL_7532487, EPI_ISL_7532636, EPI_ISL_7532667, EPI_ISL_7533126, EPI_ISL_7533558, EPI_ISL_7533573, EPI_ISL_7535208, EPI_ISL_7535261, EPI_ISL_7535308, EPI_ISL_7535330, EPI_ISL_7535331, EPI_ISL_7535381, EPI_ISL_7535476, EPI_ISL_7535510, EPI_ISL_7535600, EPI_ISL_7535650, EPI_ISL_7536373, EPI_ISL_7536571, EPI_ISL_7536608, EPI_ISL_7536664, EPI_ISL_7536712, EPI_ISL_7536778, EPI_ISL_7536955, EPI_ISL_7537157, EPI_ISL_7537194, EPI_ISL_7537382, EPI_ISL_7537419, EPI_ISL_7537698, EPI_ISL_7537754, EPI_ISL_7537774, EPI_ISL_7537864, EPI_ISL_7538259, EPI_ISL_7538311, EPI_ISL_7538654, EPI_ISL_7538855, EPI_ISL_7539129, EPI_ISL_7539442, EPI_ISL_7541246, EPI_ISL_7541250, EPI_ISL_7541383, EPI_ISL_7541410, EPI_ISL_7541455, EPI_ISL_7541551, EPI_ISL_7541700, EPI_ISL_7541743, EPI_ISL_7541804, EPI_ISL_7541826, EPI_ISL_7541859, EPI_ISL_7541865, EPI_ISL_7541886, EPI_ISL_7541892, EPI_ISL_7541941, EPI_ISL_7541961, EPI_ISL_7541994 | | | |
| see above | Lighthouse Lab in Milton Keynes | Wellcome Sanger Institute for the COVID-19 Genomics UK (COG-UK) Consortium | Cordelia Langford; David K. Jackson; Dominic Kwiatkowski; Ewan Harrison; Ian Johnston; Jeffrey Barrett; John Sillitoe on behalf of the Wellcome Sanger Institute COVID-19 Surveillance Team; Roberto Amato; Sonia Goncalves; The Lighthouse Lab in Milton Keynes and Alex Alderton |
| EPI_ISL_7201997, EPI_ISL_7302326, EPI_ISL_7302955, EPI_ISL_7303330 | Lighthouse Laboratory Plymouth | Wellcome Sanger Institute for the COVID-19 Genomics UK (COG-UK) Consortium | Cordelia Langford; David K. Jackson; Dominic Kwiatkowski; Ewan Harrison; Ian Johnston; Jeffrey Barrett; John Sillitoe on behalf of the Wellcome Sanger Institute COVID-19 Surveillance Team; Lighthouse Laboratory Plymouth and Alex Alderton; Roberto Amato; Sonia Goncalves |
| EPI_ISL_7413422, EPI_ISL_7413427, EPI_ISL_7413439, EPI_ISL_7413450 | MB-Cadham Provincial laboratory | National Microbiology Laboratory (NML) | Anna Majer; Anneliese Landgraff; CanCOGeN's metadata curation team; Darian Hole; David Alexander; Elsie Grudeski; Gary Van Domselaar; Grace Seo; Jared Bullard; Jennifer Tanner; Kerry Dust; Kirsten Biggar; Madison Chapel; Morag Graham; Natalie Knox; Nathalie Bastien; Paul Van Caeseele; Philip Mabon; Public Health Agency of Canada CanCOGeN team; Rhiannon Huzarewich; Russell Mandes; Shari Tyson; Timothy Booth; Yan Li |
| EPI_ISL_7405692 | MCL Medizinische Laboratorien, Hauptstandort Niederwangen | Institute for Infectious Diseases, University of Bern | Alban Ramette; Christian Baumann; Cora Sägesser; Franziska Suter-Riniker; Loïc Borcard; Miguel A Terrazos Miani; Nicole Liechti; Pascal Bittel; Peter Keller; Sonja Gempeler; Stefan Neuenschwander; Stephen L Leib |
| EPI_ISL_7444012, EPI_ISL_7444038, EPI_ISL_7444057 | MVZ Labor Dr. Fenner und Kollegen (Standort Hamburg) | Robert Koch Institute | |
| EPI_ISL_7418337 | MVZ Labor Dr. Limbach & Kollegen GbR | Robert Koch Institute | |
| EPI_ISL_7542261 | MVZ Rheumatologie und | Heinrich Pette Institute, Leibniz | Adam Grundhoff; Alexis Robitaille; Johannes Knobloch; Martin Aepfelbacher; Nicole Fischer; Thomas Günther |

| | | | |
|---|---|---|---|
| | Autoimmunmedizin Hamburg GmbH | Institute for Experimental Virology | |
| EPI_ISL_7154400, EPI_ISL_7154401, EPI_ISL_7154402, EPI_ISL_7500444 | Mako Medical | Centers for Disease Control and Prevention Division of Viral Diseases, Pathogen Discovery | Benjamin Rambo-Martin; Christopher Gulvick; Clinton Paden; Dakota Howard; Dhwani Batra; Duncan MacCannell; Erisa Sula; Jason Caravas; Kristine Lacek; Lauren Moon; Matthew Schmerer; Matthew Tugwell; Peter Cook; Scott Sammons; Shatavia Morrison; Tymeckia Kendall; Victoria Caban Figueroa; Yvette Unoarumhi |
| EPI_ISL_7464406, EPI_ISL_7464407, EPI_ISL_7464408 | Malawi Liverpool Wellcome Trust Clinical Research Program | Malawi Liverpool Wellcome Trust Clinical Research Program | Belson Kutambe; Ben Morton; Catherine Anscombe; Kondwani Jambo; Mavis Menyere; Philip Ashton; Sam Lissauer |
| EPI_ISL_7404794 | Maryland Genomics, Institute for Genome Sciences, University of Maryland School of Medicine | Maryland Genomics, Institute for Genome Sciences, University of Maryland School of Medicine | Claire M; Fraser; George; Hazen; Holly; Humphrys; Jacques; Jain; Kevin; Kranthi; Lisa D; Luke J; Mike; Ott; Ravel; Regan; Roussey; Sadzewicz; Sandra; Tallon; Tracy; Vavikolanu |
| EPI_ISL_7469697 | Mass General Brigham | Mass General Brigham | A.E.; Adams, G.; Anahtar, M.; B.L.; B.W.; Bauer, M.; Birren; Branda, J.; Carter, A.; Cerrato, F.; Chaluvadi, S.; Chapman; Cusick, C.; D.J.; DeRuff, K.; E. and Sabeti; Flowers, K.; Gallagher, G.; Gladden-Young, A.; Gnirke, A.; Harris, J.; J.E.; K.J.; LaRocque, R.; Lagerborg, K.; Lemieux; Lin; Loreth, C.; MacInnis; Neumann, A.; Normandin, E.; P.C.; Park; Pierce, V.; Reilly, S.; Rosenberg; Rudy, M.; Ryan, E.; S.B.; Shaw, B.; Siddle; Slater, D.; Smole, S.; Tomkins-Tinch, C.; Turbett, S.; Uddin, R. |
| EPI_ISL_6886593, EPI_ISL_6886594, EPI_ISL_6886595, EPI_ISL_6886596 | Max von Pettenkofer Institute, Virology, National Reference Center for Retroviruses, LMU Munich | Laboratory for Functional Genome Analysis; Dept. Genomics; Gene Center of the LMU Munich | Alexander Graf; Helmut Blum; Max Muenchhoff; Oliver Keppler; Stefan Krebs |
| EPI_ISL_7267259, EPI_ISL_7470216, EPI_ISL_7470264, EPI_ISL_7470331 | Medical Microbiology Unit, Department for Laboratory Medicine, Drammen Hospital, Vestre Viken Health Trust | Norwegian Institute of Public Health, Department of Virology | Atiya R Ali; Debech Nadia; Engebretsen Serina Beate; Garcia Llorente Ignacio; Hilde Elshaug; Hilde Vollan; Jon Bråte; Kamilla Heddeland Instefjord; Karoline Bragstad; Kathrine Stene-Johansen; Line Victoria Moen; Marie Paulsen Madsen; Olav Hungnes; Pedersen Benedikte Nevjen; Rasmus Riis Kopperud |
| EPI_ISL_7443804, EPI_ISL_7443805, EPI_ISL_7443809, EPI_ISL_7443815 | Medizinische Laboratorien Düsseldorf | Robert Koch Institute | |
| EPI_ISL_7416680 | Medizinisches Versorgungszentrum für Labormedizin und Mikrobiologie Ruhr GmbH - mvzlm RUHR GmbH | Robert Koch Institute | |
| EPI_ISL_6854346, EPI_ISL_6854347, EPI_ISL_6854348 | Microbiologia e Virologia Cotugno | Microbiologia e Virologia Cotugno | Antonio Canonico; Antonio Fascione; Claudia Tiberio; Enza Mallardo; Francesco Nappo; Giovanni D'Auria; Giuseppe di Gennaro; Ilaria Cavallaro; Luigi Atripaldi |
| EPI_ISL_7462324 | Microbiology Department, University Hospital Araba | Microbiology Department, University Hospital Donostia | Cilla G.; Gomez M; Hernaez S; Marimon JM; Martin-Peñaranda T; Montes M; Piñeiro L; Sorarrain A |
| EPI_ISL_7502103, EPI_ISL_7502107 | Microbiology Department. Complexo Hospitalario Universitario de Vigo | Microbiology Department. Complexo Hospitalario Universitario de Vigo | Alvarez M; Cabrera JJ; Carballo R; Cortizo S; Davina C; Martinez L; Mediero G; Pena I; Perez S; Potel C; Regueiro B; Rey S; Vasallo FJ; del-Campo V |
| EPI_ISL_7496678 | Minnesota Department of Health, Public Health Laboratory | Minnesota Department of Health, Public Health Laboratory | Alyssa Mondelli; Elizabeth Horn; Jacob Garfin; Kelly Pung; Matt Plumb; Sarah Namugenyi; and Xiong Wang |
| EPI_ISL_6963002 | Mirialis | CNR Virus des Infections Respiratoires - France SUD | Antoine Oblette; Antonin Bal; Bruno Lina; Bruno Simon; Camille Delcloitre; Eva Oddoux; Florence Morfin; Gregory Destras; Gwendolyne Burfin; Hadrien Regue; Hervé Crehalet; Jean François Bore; Jeremy Cordier; Laurence Josset; Martine Valette; Noémie Fessy; Quentin Semanas; Richard Chalvignac; Thibault Corsin; Thibault Gouiran |
| EPI_ISL_7193991, EPI_ISL_7286479, EPI_ISL_7516233, EPI_ISL_7519931, EPI_ISL_7527590, EPI_ISL_7527959 | Molekylær Medicinsk Afdeling, Aarhus University Hospital, Aarhus, Denmark | Statens Serum Institut Bioinformatics and Microbial Genomics | Danish Covid-19 Genome Consortium |
| EPI_ISL_7501186 | Mount Auburn Hospital via Lahey Hospital | New England Biolabs | Abel, G.; B.W.; C.J.; Colgrove, R.; Duncan, R.; Elfahal, M.; Flynn; Heim, K.; Karolides, M.; L. and Langhorst; Michaels, L.; Pinet, K.; Skelton, T.; Sun |
| EPI_ISL_7545652, EPI_ISL_7545653, EPI_ISL_7545654, EPI_ISL_7545658, EPI_ISL_7545660, EPI_ISL_7545661, EPI_ISL_7545662, EPI_ISL_7545663, EPI_ISL_7545664, EPI_ISL_7545665, EPI_ISL_7545666, EPI_ISL_7545667, EPI_ISL_7545668, EPI_ISL_7545669, EPI_ISL_7545670, EPI_ISL_7545671, EPI_ISL_7545673, EPI_ISL_7545674, EPI_ISL_7545675 | | | |
| see above | NHLS Livingstone Laboratory | CERI, Centre for Epidemic Response and Innvoation, Stellenbosch University and KRISP, KZN Research Innovation and Sequencing Platform, UKZN. | Arisha Maharaj; Giandhari J; Naidoo Y; Oluwakemi Laguda-Akingba and Nokukhanya Mdlalose; Pillay S; Ramphal U; Ramphal Y; San JE; Tegally H; Tshiabuila D; Wilkinson E; de Oliveira T |
| EPI_ISL_7381208, EPI_ISL_7381209, EPI_ISL_7381210, EPI_ISL_7381211, EPI_ISL_7381212, EPI_ISL_7381213, EPI_ISL_7381214, EPI_ISL_7381215, EPI_ISL_7381216, EPI_ISL_7381217, EPI_ISL_7381218, EPI_ISL_7381219, EPI_ISL_7381220, EPI_ISL_7381221, EPI_ISL_7381222 | | | |
| see above | NHLS Port Elizabeth Laboratory | CERI, Centre for Epidemic Response and Innvoation, Stellenbosch University and KRISP, KZN Research Innovation and Sequencing Platform, UKZN. | Arisha Maharaj; Giandhari J; Moir M; Naidoo Y; Oluwakemi Laguda-Akingba and Nokukhanya Mdlalose; Pillay S; Ramphal U; Ramphal Y; San JE; Tegally H; Tshiabuila D; Wilkinson E; de Oliveira T; van Wyk S |
| EPI_ISL_7462390, EPI_ISL_7462391, EPI_ISL_7462392, EPI_ISL_7462393, EPI_ISL_7462397, EPI_ISL_7462398, EPI_ISL_7462399, EPI_ISL_7462400, EPI_ISL_7462401, EPI_ISL_7462402, EPI_ISL_7462403, EPI_ISL_7462404 | | | |
| see above | NHLS Universitas Academic | Division of Medical Virology, National Health Laboratory Service (NHLS), Tygerberg Hospital / Stellenbosch University | D Goedhals; Emmanuel Ogunbayo; MM Nyaga; MT Mogotsi; P Nthiga; PA Bester; Shannon Wilson; Susan Engelbrecht; T de Oliveira; Tongai Maponga; Wolfgang Preiser |
| EPI_ISL_7285023 | National Centre for Disease Control (NCDC) Biotechnology Division, Delhi | NCDC Delhi, Biotechnology Division INSACOG | Hema Gogia; Hemlata Lall; Kalaiarasan Ponnusamy; Mahesh S Dhar; Manoj K Singh; Meena Datta; Partha Rakshit; Preeti Madan; Priyanka Singh; Radhakrishnan V. S; Robin Marwal; Sandhya Kabra; Sujeet K Singh; Uma Sharma |
| EPI_ISL_7548907, EPI_ISL_7548908, EPI_ISL_7548911, EPI_ISL_7548912, EPI_ISL_7548913, EPI_ISL_7548915, EPI_ISL_7548916, EPI_ISL_7548918, EPI_ISL_7548919, EPI_ISL_7548920, EPI_ISL_7548921, EPI_ISL_7548922, EPI_ISL_7548924, EPI_ISL_7548925, EPI_ISL_7548926, EPI_ISL_7548927, EPI_ISL_7548928, EPI_ISL_7548929, EPI_ISL_7548930, EPI_ISL_7548932, EPI_ISL_7548933, EPI_ISL_7548934, EPI_ISL_7548935, EPI_ISL_7548936, EPI_ISL_7548937, EPI_ISL_7548938, EPI_ISL_7548939 | | | |
| see above | National Health Laboratory | Botswana Harvard AIDS Institute Partnership, Plot 1836 North Ring Road, Princess Marina Hospital, Gaborone | Boitumelo Zuze; Botshelo Radibe; Dorcas Maruapula; Doreen Ditshwanelo; Joseph Makhema; Keoratile Ntshambiwa; Kgomotso Moruisi; Legodile Kooepile; Mosepele Mosepele; Mphaphi B. Mbulawa; Ontlametse T. Bareng; Pamela Smith-Lawrence; Roger Shapiro; Sefetogi Ramaologa; Shahin Lockman; Sikhulile Moyo; Simani Gaseitsiwe; Thongbotho Mphoyakgosi; Wonderful T. Choga |
| EPI_ISL_6795195, EPI_ISL_6795199, EPI_ISL_6795202, EPI_ISL_6795203, EPI_ISL_6795406, EPI_ISL_7015210, EPI_ISL_7015212, EPI_ISL_7015216, EPI_ISL_7015223, EPI_ISL_7015224, EPI_ISL_7015226, EPI_ISL_7015229, EPI_ISL_7015230, EPI_ISL_7310589, EPI_ISL_7310595, EPI_ISL_7310605, EPI_ISL_7310613, EPI_ISL_7310622, EPI_ISL_7310630, EPI_ISL_7310631, EPI_ISL_7310636, EPI_ISL_7310643, EPI_ISL_7310648, EPI_ISL_7310658, EPI_ISL_7310666, EPI_ISL_7310675, EPI_ISL_7310679, EPI_ISL_7310685, EPI_ISL_7310703, EPI_ISL_7310710, EPI_ISL_7310719, EPI_ISL_7310725, EPI_ISL_7310733, EPI_ISL_7310742, EPI_ISL_7310747, EPI_ISL_7358050, EPI_ISL_7358051, EPI_ISL_7358052, EPI_ISL_7358053, EPI_ISL_7358054, EPI_ISL_7358055, EPI_ISL_7358056, EPI_ISL_7358057, EPI_ISL_7358058, EPI_ISL_7358059, EPI_ISL_7358064, EPI_ISL_7358065, EPI_ISL_7358066, EPI_ISL_7358067, EPI_ISL_7358068, EPI_ISL_7358069, EPI_ISL_7358070, EPI_ISL_7358071, EPI_ISL_7358072, EPI_ISL_7358073, EPI_ISL_7358074, EPI_ISL_7358075, EPI_ISL_7358076, EPI_ISL_7358077, EPI_ISL_7358078, EPI_ISL_7358079, EPI_ISL_7358080, EPI_ISL_7358081, EPI_ISL_7358082, EPI_ISL_7358083, EPI_ISL_7358084, EPI_ISL_7358085, EPI_ISL_7358089, EPI_ISL_7358090, EPI_ISL_7358091, EPI_ISL_7358092, EPI_ISL_7358093, EPI_ISL_7381192, EPI_ISL_7381193, EPI_ISL_7381194, EPI_ISL_7381195, EPI_ISL_7381196, EPI_ISL_7381197, EPI_ISL_7381198, EPI_ISL_7381199, EPI_ISL_7381200, EPI_ISL_7381201, EPI_ISL_7381202, EPI_ISL_7381203, EPI_ISL_7381204, EPI_ISL_7381205, EPI_ISL_7381206, EPI_ISL_7381207, EPI_ISL_7545676, EPI_ISL_7545677, EPI_ISL_7545678, EPI_ISL_7545679, EPI_ISL_7545680, EPI_ISL_7545681, EPI_ISL_7545682, EPI_ISL_7545683, EPI_ISL_7545684, EPI_ISL_7545685, EPI_ISL_7545686, EPI_ISL_7545687, EPI_ISL_7545688, EPI_ISL_7545689, EPI_ISL_7545690, EPI_ISL_7545691, EPI_ISL_7545692, EPI_ISL_7545693, EPI_ISL_7545694, EPI_ISL_7545695, EPI_ISL_7545696, EPI_ISL_7545698, EPI_ISL_7545699, EPI_ISL_7545700, EPI_ISL_7545701, EPI_ISL_7545702, EPI_ISL_7545703, EPI_ISL_7545704, EPI_ISL_7545705, EPI_ISL_7545706, EPI_ISL_7545707, EPI_ISL_7545708, EPI_ISL_7545709, EPI_ISL_7545710, EPI_ISL_7545711, EPI_ISL_7545712, EPI_ISL_7545713, EPI_ISL_7545714, EPI_ISL_7545715, EPI_ISL_7545716, EPI_ISL_7545717, EPI_ISL_7545718, EPI_ISL_7545719, EPI_ISL_7545720, EPI_ISL_7545721, EPI_ISL_7545722, EPI_ISL_7545723, EPI_ISL_7545724, EPI_ISL_7545725, EPI_ISL_7545726, EPI_ISL_7545727, EPI_ISL_7545728, EPI_ISL_7545729, EPI_ISL_7545730, EPI_ISL_7545731, EPI_ISL_7545732, EPI_ISL_7545733, EPI_ISL_7545734, EPI_ISL_7545735, EPI_ISL_7545736, EPI_ISL_7545737, EPI_ISL_7545738, EPI_ISL_7545739, EPI_ISL_7545742, EPI_ISL_7545743, EPI_ISL_7545744, EPI_ISL_7545745, EPI_ISL_7545746, EPI_ISL_7545747, EPI_ISL_7545748, EPI_ISL_7545749, EPI_ISL_7545750, EPI_ISL_7545751, EPI_ISL_7545752, EPI_ISL_7545753, EPI_ISL_7545754, EPI_ISL_7545755, EPI_ISL_7545756, EPI_ISL_7545757, EPI_ISL_7545758, EPI_ISL_7545759, EPI_ISL_7545760, EPI_ISL_7545761, EPI_ISL_7545762, EPI_ISL_7545764, EPI_ISL_7545765, EPI_ISL_7545766, EPI_ISL_7545767, EPI_ISL_7545768, EPI_ISL_7545769, EPI_ISL_7545770, EPI_ISL_7545771, EPI_ISL_7545772, EPI_ISL_7545773, EPI_ISL_7545774, EPI_ISL_7545775, EPI_ISL_7545776, EPI_ISL_7545777, EPI_ISL_7545778, EPI_ISL_7545779, EPI_ISL_7545780, EPI_ISL_7545781, EPI_ISL_7545783, EPI_ISL_7545784, EPI_ISL_7545785, EPI_ISL_7545787, EPI_ISL_7545788, EPI_ISL_7545789, EPI_ISL_7545790, EPI_ISL_7545791, EPI_ISL_7545792, EPI_ISL_7545793, EPI_ISL_7545794 | | | |
| see above | National Health Laboratory Service, Kwazulu-Natal, South Africa | CERI, Centre for Epidemic Response and Innvoation, Stellenbosch University and KRISP, KZN Research Innovation and Sequencing Platform, UKZN. | Arisha Maharaj; Giandhari J; Moir M; Naidoo Y; Nokukhanya Mdlalose; Pillay S; Ramphal U; Ramphal Y; San JE; Tegally H; Tshiabuila D; Wilkinson E; de Oliveira T; van Wyk S |
| EPI_ISL_6829557 | National Health Laboratory Service, Kwazulu-Natal, South Africa | KRISP, KZn Research Innovation and Sequencing Platform | Arisha Maharaj; Giandhari J; Lessells R; Moir M; Naidoo Y; Nokukhanya M; Pillay S; Ramphal U; Ramphal Y; San JE; Tegally H; Tshiabuila D; Wilkinson E; de Oliveira T |
| EPI_ISL_6795188, EPI_ISL_6795189, EPI_ISL_6795190, EPI_ISL_6795191, EPI_ISL_6795192, EPI_ISL_6795193 | National Health Laboratory Services, Virology | CERI, Centre for Epidemic Response and Innvoation, Stellenbosch University and KRISP, KZN Research Innovation and Sequencing Platform, UKZN. | Arisha Maharaj; Florette Treurnicht; Giandhari J; Kathleen Subramoney; Naidoo Y; Pillay S; Ramphal U; Ramphal Y; San JE; Tegally H; Tshiabuila D; Wilkinson E; de Oliveira T |
| EPI_ISL_6699728, EPI_ISL_6699729, EPI_ISL_6699730, EPI_ISL_6699731, EPI_ISL_6699732, EPI_ISL_6699733, EPI_ISL_6699734, EPI_ISL_6699735, EPI_ISL_6699736, EPI_ISL_6699737, EPI_ISL_6699738, EPI_ISL_6699739, EPI_ISL_6699740, EPI_ISL_6699741, EPI_ISL_6699742, EPI_ISL_6699743, EPI_ISL_6699744, EPI_ISL_6699745, EPI_ISL_6699746, EPI_ISL_6699747, EPI_ISL_6699748, EPI_ISL_6699749, EPI_ISL_6699750, EPI_ISL_6699751, EPI_ISL_6699752, EPI_ISL_6699753, EPI_ISL_6699754, EPI_ISL_6699755, EPI_ISL_6699756, EPI_ISL_6699757, EPI_ISL_6699758, EPI_ISL_6699759, EPI_ISL_6699760, EPI_ISL_6699761, EPI_ISL_6699762, EPI_ISL_6699763, EPI_ISL_6699764, EPI_ISL_6699765, EPI_ISL_6699766, EPI_ISL_6699767, EPI_ISL_6699768, EPI_ISL_6699769, EPI_ISL_6699770, EPI_ISL_6699771, EPI_ISL_6782043, EPI_ISL_6782055, EPI_ISL_6782056, EPI_ISL_6782071, EPI_ISL_6782079, EPI_ISL_6782080, EPI_ISL_6782084, EPI_ISL_6782090, EPI_ISL_6782092, EPI_ISL_6810482, EPI_ISL_6810483, EPI_ISL_6810484, EPI_ISL_6810485, EPI_ISL_6810486, EPI_ISL_6810487 | | | |

| | | | |
|---|---|---|---|
| see above | National Health Laboratory Services, Virology, Charlotte Maxeke Johannesburg hospital, Parktown, Johannesburg, Gauteng | CERI, Centre for Epidemic Response and Innvoation, Stellenbosch University and KRISP, KZN Research Innovation and Sequencing Platform, UKZN. | Amoaka D; Arisha Maharaj; Avani Bharuthram; Bester P; Bhiman J; Engelbrecht S; Everatt J; Florette Treurnicht; Goedhals D; Hardie D; Hsiao M; Iranzadeh A; Kathleen Subramoney; Lessells R; Makatini Z; Maponga T; Mdlalose N; Mlisana K; Moir M; NGS-SA (Scheepers C; Naidoo Y; Nkhensani Mtileni; Nyaga M) Giandhari J; Oluwakemi M; Pillay S; Preiser W; Ramphal U; Ramphal Y; San JE; Tegally H; Tshiabuila D; Venter M; Wilkinson E; Williamson C; de Oliveira T; von Gottberg A |
| EPI_ISL_6939033, EPI_ISL_6939034, EPI_ISL_6939035, EPI_ISL_6939036, EPI_ISL_6939038, EPI_ISL_6939039, EPI_ISL_6939041, EPI_ISL_6939042, EPI_ISL_6939043, EPI_ISL_6939044, EPI_ISL_6939045, EPI_ISL_6939046, EPI_ISL_6939047, EPI_ISL_6939048, EPI_ISL_6939049, EPI_ISL_6939050, EPI_ISL_6939051, EPI_ISL_6939052, EPI_ISL_6939053, EPI_ISL_6939054, EPI_ISL_6939056, EPI_ISL_6939057, EPI_ISL_6939058, EPI_ISL_6939059, EPI_ISL_6939060, EPI_ISL_6939061, EPI_ISL_6939062, EPI_ISL_6939063, EPI_ISL_6939064, EPI_ISL_6939065, EPI_ISL_6939066, EPI_ISL_6939067, EPI_ISL_6939068 | | | |
| see above | National Influenza Centre | National Influenza Centre | ; Benjamiin B. Lindsey; Benjamin H. Foulkes; Bless Seyram Agbenyo; Bright Adu; Ernest Asiedu; Franklin Asiedu-Bekoe; Hilda Opoku Frempong; Ivy A. Asante; Joseph Oliver-Commey; Joyce Appiah-Kubi; Keren Okyerebea Attiku; Linda Boatemaa; Lorreta Kwasah; Mathew D. Parker; Michael Marks; Mildred Adusei-Poku; Quaneeta Mohktar; Sharon Hsu; Thushan I de Silva; William K. Ampofo |
| EPI_ISL_7456448, EPI_ISL_7456450, EPI_ISL_7456451, EPI_ISL_7456452, EPI_ISL_7456453, EPI_ISL_7456454, EPI_ISL_7456455, EPI_ISL_7456456, EPI_ISL_7456457 | | | |
| see above | National Institute For Communicable Diseases Of The National Health Laboratory Service | National Institute for Communicable Diseases of the National Health Laboratory Service | Amoako DG; Bhiman JN; Everatt J; Ismail A; Mahlangu B; Mnguni A; Mohale T; Ntuli N; Scheepers C; Wolter N |
| EPI_ISL_7418017 | National Institute of Infectious Diseases | National Institute of Infectious Diseases | Harutaka Katano; Ken Maeda; Kentaro Itokawa; Makoto Kuroda; Shuetsu Fukushi; Shun Iida; Tadaki Suzuki; Tsuyoshi Sekizuka; Yudai Kuroda |
| EPI_ISL_7021517, EPI_ISL_7074135 | National Platform bis COVID ULB-IBC | National Platform bis COVID ULB-IBC | Arnaud Marchant; Benoit Haerlingen; Coralie Henin; Marie-Luce Delforge; Ricardo De Mendonça |
| EPI_ISL_7063764, EPI_ISL_7288348, EPI_ISL_7288357, EPI_ISL_7288375, EPI_ISL_7544936, EPI_ISL_7544937, EPI_ISL_7544938, EPI_ISL_7544939, EPI_ISL_7544940, EPI_ISL_7544941, EPI_ISL_7544942, EPI_ISL_7544947, EPI_ISL_7544948, EPI_ISL_7544949, EPI_ISL_7544950, EPI_ISL_7544953, EPI_ISL_7544954, EPI_ISL_7544956, EPI_ISL_7544960, EPI_ISL_7544963, EPI_ISL_7544964, EPI_ISL_7544965, EPI_ISL_7544969, EPI_ISL_7544971, EPI_ISL_7544972, EPI_ISL_7544974, EPI_ISL_7544975, EPI_ISL_7544980 | | | |
| see above | National Platform bis UMONS/Jolimont | National Platform bis UMONS/Jolimont | Eric Tarantino; Florian Juszczak; Gautier Detry; Guillaume Bayon-Vicente; Laetitia Gheysen; Ruddy Wattiez |
| EPI_ISL_7137310, EPI_ISL_7137311, EPI_ISL_7195620, EPI_ISL_7195621, EPI_ISL_7195622, EPI_ISL_7195623, EPI_ISL_7460338 | | | |
| see above | National Public Health Laboratory, National Centre for Infectious Diseases | National Public Health Laboratory, National Centre for Infectious Diseases | Benny Yeo Ken Yee; Constance Chen; Dennis Loy Song Qi; Dimitar Kenanov; Grace Ngan Jie Yin; Katherine Ching Zi Yan; Kwan Ki Ko; Lin Cui; Mak Tze Minn; Niranjan Nagarajan; Raymond Tzer Pin Lin; Royce Ang; Samuel Loo; Sebastian Maurer Stroh; Suphavilai Chayaporn; Zhenyang Zhou |
| EPI_ISL_7154394, EPI_ISL_7154403, EPI_ISL_7273097, EPI_ISL_7547734, EPI_ISL_7547735 | National Reference Laboratory, NCDC | National Reference Laboratory, Nigeria Centre for Disease Control | Catherine Okoi; Chimaobi Chukwu; Dr Ifedayo Adetifa; Dr Ndodo Nnaemeka; Dr Omoare Adesuyi; Nwando Mba; Olajumoke Babatunde; Olusola Anuoluwapo Akanbi; Oyeronke Ayansola |
| EPI_ISL_6939819, EPI_ISL_7354124, EPI_ISL_7406048, EPI_ISL_7406049, EPI_ISL_7406082, EPI_ISL_7406083 | National Virus Reference Laboratory | National Virus Reference Laboratory | Charlene Bennett; Cillian F De Gascun; Gabriel Gonzalez; Jonathan Dean; Michael Carr; Zoe Yandle |
| EPI_ISL_7116413, EPI_ISL_7116439, EPI_ISL_7116461, EPI_ISL_7116681, EPI_ISL_7116718, EPI_ISL_7116738 | Nebraska Public Health Laboratory | NPHL COVID-19 Response Team | NPHL COVID-19 Response Team |
| EPI_ISL_7544379, EPI_ISL_7544739 | New Somerset Hospital wc NSH | NHLS/UCT | Arash Iranzadeh; Bruna Galvao; Carolyn Williamson; Deelan Doolabh; Diana Hardie; Gert Marais; Innocent Mudau; Luicer Olubayo; Lynn Tyers; Marvin Hsiao; Nokuzola Mbhele; Rageema Joseph; Stephen Korsman |
| EPI_ISL_6814922, EPI_ISL_6814923, EPI_ISL_6829575, EPI_ISL_6829577, EPI_ISL_6958955, EPI_ISL_7162071, EPI_ISL_7162072, EPI_ISL_7162073, EPI_ISL_7265843, EPI_ISL_7379517, EPI_ISL_7379527, EPI_ISL_7457536, EPI_ISL_7457537, EPI_ISL_7457538, EPI_ISL_7503742, EPI_ISL_7503743, EPI_ISL_7503744, EPI_ISL_7503745, EPI_ISL_7503746 | | | |
| see above | New South Wales Health Pathology Royal Prince Alfred Hospital | Microbiology RPAH | Au, J.; Bull, R.; Deveson, I.; Foster, C.; Rawlinson, W.; Ruiz Silva, M.; Van Hal, S. |
| EPI_ISL_7418387, EPI_ISL_7431953 | Niedersächsisches Landesgesundheitsamt (NLGA) | Robert Koch Institute | |
| EPI_ISL_6958280 | North Lantau Hospital | Hong Kong Department of Health | Alan K.L. Tsang; Edman T.K. Lam; Ken H.L. Ng; Peter C.W. Yip; Rickjason C.W. Chan |
| EPI_ISL_7334887, EPI_ISL_7334888 | ONEIDA COUNTY HEALTH DEPT. | Wadsworth Center, New York State Department of Health | Alexis Russell; Catharine Prussing; Daryl M. Lamson; Erasmus Schneider; Erica Lasek-Nesselquist; John Kelly; Jonathan Plitnick; Kirsten St. George; Matthew Shudt; Melissa A Leisner; Navjot Singh |
| EPI_ISL_7495763 | Omega Diagnostics at Mounes | Omega Diagnostics at Mounes | Cherish Jackson; Cynthia Corley; Latira Haynes-Jacob; MD; Vivek Khare |
| EPI_ISL_7040235 | Oslo University Hospital, Department of Medical Microbiology | Norwegian Institute of Public Health, Department of Virology | Atiya R Ali; Debech Nadia; Engebretsen Serina Beate; Garcia Llorente Ignacio; Hilde Elshaug; Hilde Vollan; Jon Bråte; Kamilla Heddeland Instefjord; Karoline Bragstad; Kathrine Stene-Johansen; Line Victoria Moen; Marie Paulsen Madsen; Olav Hungnes; Pedersen Benedikte Nevjen; Rasmus Riis Kopperud |
| EPI_ISL_7248730, EPI_ISL_7248745, EPI_ISL_7248753, EPI_ISL_7248763, EPI_ISL_7470724, EPI_ISL_7470728 | Oslo University Hospital, Department of Microbiology | Norwegian Institute of Public Health, Department of Virology | Arvind Yegambaram Meenakshi Sundaram; Atiya R Ali; Cathrine Fladeby; Debech Nadia; Engebretsen Serina Beate; Garcia Llorente Ignacio; Gregor D. Gilfillan; Hilde Elshaug; Hilde Vollan; Jon Bråte; Kamilla Heddeland Instefjord; Karoline Bragstad; Kathrine Stene-Johansen; Line Victoria Moen; Lise Andresen; Mariann Nilsen; Marie Paulsen Madsen; Mona Holberg-Petersen; Olav Hungnes; Pedersen Benedikte Nevjen; Pål Marius Bjørnstad; Rasmus Riis Kopperud; Teodora Plamenova Ribarska |
| EPI_ISL_7470189 | Ostfold Hospital Trust - Kalnes, Centre for Laboratory Medicine, Section for gene technology and infection serology | Norwegian Institute of Public Health, Department of Virology | Atiya R Ali; Debech Nadia; Engebretsen Serina Beate; Garcia Llorente Ignacio; Hilde Elshaug; Hilde Vollan; Jon Bråte; Kamilla Heddeland Instefjord; Karoline Bragstad; Kathrine Stene-Johansen; Line Victoria Moen; Marie Paulsen Madsen; Olav Hungnes; Pedersen Benedikte Nevjen; Rasmus Riis Kopperud |
| EPI_ISL_7467356 | PHV-FSS | PHV-FSS | Chenwei Wang on behalf of Q-PHIRE Genomics |
| EPI_ISL_6774082, EPI_ISL_6774086, EPI_ISL_6774092 | Palapye Primary Hospital Laboratory | Botswana Harvard HIV Reference Laboratory | Boitumelo Zuze; Botshelo Radibe; Dorcas Maruapula; Joseph Makhema; Keoratile Ntshambiwa; Kgomotso Moruisi; Legodile Kooepile; Mosepele Mosepele; Mphaphi B. Mbulawa; Ontlametse T. Bareng; Pamela Smith-Lawrence; Roger Shapiro; Sefetogi Ramaologa; Shahin Lockman; Sikhulile Moyo; Simani Gaseitsiwe; Thongbotho Mphoyakgosi; Wonderful T. Choga |
| EPI_ISL_7129868, EPI_ISL_7129869 | Pandemic Response Lab - NYC | Pandemic Response Lab, R&D | Alex Carpio; Cybill del Castillo; Dylan Law; Haiping Hao; Henry Lee; Isabel Fernandez Escapa; Jon Laurent; Melissa Hopkins; Michael Hammerling; Pradeep Bugga; Shinyoung Clair Kang; Sol Rey; William Ward |
| EPI_ISL_6842152, EPI_ISL_6842154, EPI_ISL_6842155, EPI_ISL_6842157, EPI_ISL_6842158, EPI_ISL_6842160, EPI_ISL_6842161, EPI_ISL_6842164, EPI_ISL_6842166, EPI_ISL_6842167, EPI_ISL_7452739, EPI_ISL_7452740, EPI_ISL_7452743, EPI_ISL_7452747, EPI_ISL_7452752, EPI_ISL_7452753, EPI_ISL_7452754, EPI_ISL_7452755, EPI_ISL_7452756, EPI_ISL_7452757, EPI_ISL_7452759, EPI_ISL_7452760, EPI_ISL_7452779, EPI_ISL_7452784, EPI_ISL_7452786, EPI_ISL_7452787, EPI_ISL_7452788, EPI_ISL_7452789, EPI_ISL_7452790, EPI_ISL_7452791, EPI_ISL_7452801, EPI_ISL_7452802, EPI_ISL_7452803, EPI_ISL_7452804, EPI_ISL_7456466, EPI_ISL_7456467, EPI_ISL_7456468, EPI_ISL_7456469, EPI_ISL_7456470, EPI_ISL_7456471, EPI_ISL_7456472, EPI_ISL_7456473, EPI_ISL_7456474, EPI_ISL_7456475, EPI_ISL_7456476, EPI_ISL_7456477, EPI_ISL_7456478, EPI_ISL_7456479, EPI_ISL_7456480, EPI_ISL_7456481, EPI_ISL_7456482, EPI_ISL_7456483, EPI_ISL_7456484, EPI_ISL_7456485, EPI_ISL_7456489, EPI_ISL_7456490, EPI_ISL_7456491, EPI_ISL_7456492, EPI_ISL_7456493, EPI_ISL_7456494, EPI_ISL_7456495, EPI_ISL_7456496, EPI_ISL_7456497, EPI_ISL_7456498, EPI_ISL_7456499, EPI_ISL_7456500, EPI_ISL_7456501, EPI_ISL_7456502, EPI_ISL_7456503, EPI_ISL_7456504, EPI_ISL_7456505, EPI_ISL_7456506, EPI_ISL_7456507, EPI_ISL_7456508, EPI_ISL_7456509, EPI_ISL_7456510, EPI_ISL_7456511, EPI_ISL_7456512, EPI_ISL_7456513, EPI_ISL_7456514, EPI_ISL_7456515, EPI_ISL_7456516, EPI_ISL_7456517, EPI_ISL_7456518, EPI_ISL_7456519, EPI_ISL_7456520, EPI_ISL_7456521, EPI_ISL_7456522, EPI_ISL_7456523, EPI_ISL_7456524, EPI_ISL_7544906, EPI_ISL_7544907, EPI_ISL_7544908, EPI_ISL_7544909, EPI_ISL_7544910, EPI_ISL_7544911, EPI_ISL_7544912, EPI_ISL_7544913, EPI_ISL_7544914, EPI_ISL_7544915, EPI_ISL_7544916, EPI_ISL_7544917, EPI_ISL_7544918, EPI_ISL_7544919, EPI_ISL_7544920, EPI_ISL_7544921, EPI_ISL_7544922, EPI_ISL_7544923, EPI_ISL_7544924, EPI_ISL_7544925, EPI_ISL_7544926, EPI_ISL_7544927, EPI_ISL_7544928 | | | |
| see above | PathCare, Cape Town | Division of Medical Virology, National Health Laboratory Service (NHLS), Tygerberg Hospital / Stellenbosch University | Gert van Zyl; Jean Maritz; Kamela Mahlakwane; Nadine Cronje; Petra Raimond; Shannon Wilson; Tania Stander; Tongai Maponga; Wolfgang Preiser |
| EPI_ISL_7263932, EPI_ISL_7263933 | Pathogenic Microorganisms Variability Laboratory | Pathogenic Microorganisms Variability Laboratory | Alexander Gintsburg; Alexander Voskoboinikov; Andrei Siniavin; Andrey Pochtovyy; Artem Tkachuk; Denis Logunov; Elena Shidlovskaya; Elizaveta Divisenko; Inna Dolzhikova; Lyudmila Vasilchenko; Nadezhda Kuznetsova; Odintsova Alina; Vladimir Gushchin |
| EPI_ISL_7265083, EPI_ISL_7265084 | Pathology North - Royal North Shore Hospital - NSW Health Pathology | NSW Health Pathology - Institute of Clinical Pathology and Medical Research; Westmead Hospital; University of Sydney | Arnott A.; Draper J.; Gall M.; Martinez E.; Rockett R.; Sintchenko V.; on behalf of ICPMR |
| EPI_ISL_6864915, EPI_ISL_6956011, EPI_ISL_6956014 | Pathology West - NSW Health Pathology | NSW Health Pathology - Institute of Clinical Pathology and Medical Research; Westmead Hospital; University of Sydney | Arnott A.; Draper J.; Gall M.; Martinez E.; Rockett R.; Sintchenko V.; on behalf of ICPMR |
| EPI_ISL_7406251, EPI_ISL_7472848, EPI_ISL_7472859, EPI_ISL_7472865 | Plateforme de testing Namuroise | Plateforme de testing Namuroise | Degosserie Jonathan; Demars Aurore; Denis Olivier; Gilliard Nicolas; Maschietto Céline; Mullier François; Nobis Chloé; Otto Gaetan |
| EPI_ISL_7173899 | Platform BIS UZA/UAntwerpen | Labo Klinische Biologie, UZA | Basil Britto Xavier; Christine Lammens; Herman Goossens; Ines Verbesselt; Jasmine Coppens; Kathleen Holemans; Marie Le Mercier; Veerle Matheeussen |

| | | | |
|---|---|---|---|
| EPI_ISL_6777160 | Policlinico San Donato | Laboratory of Clinical Microbiology, Virology and Bioemergencies, ASST Fatebenefratelli Sacco - Sacco Hospital | Valeria Micheli |
| EPI_ISL_7416687, EPI_ISL_7416708 | Procomcure Biotech Germany GmbH | Robert Koch Institute | |
| EPI_ISL_7507116, EPI_ISL_7507117, EPI_ISL_7507119 | Public Health Authority of the Slovak Republic | Public Health Authority of the Slovak Republic | Anna Gičová; Barbora Kotvasová; Elena Tichá; Lucia Ševčíková; Miroslav Böhmer; Pavol Mišenko; Terézia Vrabľová; Tomáš Szemes |
| EPI_ISL_7135501, EPI_ISL_7135502, EPI_ISL_7135503, EPI_ISL_7135504, EPI_ISL_7263924, EPI_ISL_7263925, EPI_ISL_7263926, EPI_ISL_7263927, EPI_ISL_7263928, EPI_ISL_7263929, EPI_ISL_7263930, EPI_ISL_7334884, EPI_ISL_7334885, EPI_ISL_7334886, EPI_ISL_7438844, EPI_ISL_7438882, EPI_ISL_7438901, EPI_ISL_7438918, EPI_ISL_7438932, EPI_ISL_7438964, EPI_ISL_7438994, EPI_ISL_7459993, EPI_ISL_7459994, EPI_ISL_7459995, EPI_ISL_7459996, EPI_ISL_7459997, EPI_ISL_7459998, EPI_ISL_7495449, EPI_ISL_7495450, EPI_ISL_7495451, EPI_ISL_7495452, EPI_ISL_7495453, EPI_ISL_7495454, EPI_ISL_7495455 | | | |
| see above | Public Health Laboratory, Public Health Service Amsterdam, The Netherlands | Department of Medical Microbiology & Infection prevention, Amsterdam University Medical Centers location AMC | Akke Cornelissen; Fokla Zorgdrager; Janke Schinkel; Jelle Koopsen; Judith den Uil; Marcel Jonges; Matthijs Welkers; Menno de Jong; Robin van Houdt; Sebastien Matamoros; Sjoerd Rebers; Sylvia Bruisten; Tjalling Leenstra and Mariken van der Lubben on behalf of the Amsterdam Regional Genomic epidemiology and Outbreak Surveillance (ARGOS) consortium |
| EPI_ISL_7135499, EPI_ISL_7259732, EPI_ISL_7259741, EPI_ISL_7259744, EPI_ISL_7334879, EPI_ISL_7334880, EPI_ISL_7464493, EPI_ISL_7464494 | | | |
| see above | Public Health Ontario Laboratory | Public Health Ontario Laboratory | Aimin Li; Alireza Eshaghi; Andre Villegas; Ashleigh Sullivan; Christine Frantz; Dean Maxwell; Esha Joshi; Jared Simpson; Jennifer L Guthrie; Jonathan B Gubbay; Karthikeyan Sivaraman; Lawrence Heisler; Matthew Watson; Michael CY Li; Michael Laszloffy; Nahuel Fittipaldi; Philip Banh; Richard de Borja; Samir N Patel; Sandeep Nagra; Sandra Zittermann; Sarah Teatero; Vanessa G Allen; Yao Chen; Yogi Sundaravadanam |
| EPI_ISL_6862005 | Regional Hospital Liberec | Regional Hospital Liberec | Iva Dolinova; Katerina Arientova; Katerina Stillerova; Martin Kracik; Tomas Zajic |
| EPI_ISL_7166216, EPI_ISL_7345201, EPI_ISL_7345221, EPI_ISL_7345322, EPI_ISL_7398995, EPI_ISL_7399058, EPI_ISL_7399073, EPI_ISL_7399078, EPI_ISL_7399512, EPI_ISL_7400550, EPI_ISL_7400551, EPI_ISL_7400555, EPI_ISL_7463952, EPI_ISL_7463953, EPI_ISL_7463956, EPI_ISL_7463961, EPI_ISL_7463968, EPI_ISL_7463969, EPI_ISL_7463972, EPI_ISL_7463975, EPI_ISL_7463979, EPI_ISL_7463997, EPI_ISL_7464020, EPI_ISL_7464030, EPI_ISL_7464048, EPI_ISL_7464049, EPI_ISL_7464059, EPI_ISL_7509826, EPI_ISL_7509828, EPI_ISL_7509830 | | | |
| see above | Respiratory Virus Unit, Microbiology Services Colindale, Public Health England | COVID-19 Genomics UK (COG-UK) Consortium | PHE Covid Sequencing Team |
| EPI_ISL_7116918 | Robert Koch-Institut ZBS1 (Zentrum für biologische Gefahren und spezielle Pathogene hochpathogene Viren) | Robert Koch Institute | |
| EPI_ISL_7142714, EPI_ISL_7303373, EPI_ISL_7485193, EPI_ISL_7519924, EPI_ISL_7536363 | Rosalind Franklin Laboratory | Wellcome Sanger Institute for the COVID-19 Genomics UK (COG-UK) Consortium | Cordelia Langford; David K. Jackson; Dominic Kwiatkowski; Donald Fraser; Ewan Harrison; Ian Johnston; Jeffrey Barrett; John Sillitoe on behalf of the Wellcome Sanger Institute COVID-19 Surveillance Team; Rob Howes; Roberto Amato; Sonia Goncalves; Suki Lee; The Rosalind Franklin Laboratory and Alex Alderton |
| EPI_ISL_7366154 | Rush University Medical Center | RIPHL at Rush University Medical Center | Alyse Kittner; Cecilia Chau; Diane Springer; Edith Perez; Felix Araujo Perez; Hannah Barbian; Joyce Houlihan; Kevin Kunstman; Laura Furtado; Marieta Hyde; Mary Hayden; Sofiya Bobrovska; Stefan Green |
| EPI_ISL_7545637, EPI_ISL_7545638, EPI_ISL_7545639, EPI_ISL_7545640, EPI_ISL_7545641, EPI_ISL_7545642, EPI_ISL_7545643, EPI_ISL_7545644, EPI_ISL_7545645, EPI_ISL_7545646, EPI_ISL_7545647, EPI_ISL_7545648, EPI_ISL_7545649, EPI_ISL_7545650 | | | |
| see above | SAMRC | CERI, Centre for Epidemic Response and Innvoation, Stellenbosch University and KRISP, KZN Research Innovation and Sequencing Platform, UKZN. | Arisha Maharaj; Giandhari J; MRC; Naidoo Y; Pillay S; Ramphal U; Ramphal Y; San JE; Tegally H; Tshiabuila D; Wilkinson E; de Oliveira T |
| EPI_ISL_6913953, EPI_ISL_6914908, EPI_ISL_7194610 | SARS-CoV-2 testing team, National Institute of Infectious Diseases | Pathogen Genomics Center, National Institute of Infectious Diseases | Hazuka Y Furihata; Kentaro Itokawa; Makoto Kuroda; Masanori Hashino; Masumichi Saito; Naomi Nojiri; Nozomu Hanaoka; Rina Tanaka; Tsuguto Fujimoto; Tsuyoshi Sekizuka |
| EPI_ISL_7458718, EPI_ISL_7458719, EPI_ISL_7458720, EPI_ISL_7458721 | SK-Roy Romanow Provincial Laboratory | Saskatchewan - Roy Romanow Provincial Laboratory (RRPL) | Alanna Senecal; Amanda Lang; Jessica Minion; Kara Loos; Keith MacKenzie; Meredith Faires; Rachel DePaulo; Roy Romanow Provincial Laboratory - Molecular Diagnostics; Ryan McDonald |
| EPI_ISL_7334889, EPI_ISL_7334890 | SUNRISE MEDICAL LABORATORIES | Wadsworth Center, New York State Department of Health | Alexis Russell; Catharine Prussing; Daryl M. Lamson; Erasmus Schneider; Erica Lasek-Nesselquist; John Kelly; Jonathan Plitnick; Kirsten St. George; Matthew Shudt; Melissa A Leisner; Navjot Singh |
| EPI_ISL_7195727 | SYNLAB | GIGA Medical Genomics | Bouchra Boujemla; Claire Gourzonès; Cécile Meex; Keith Durkin; Laurent Gillet; Maria Artesi; Marie-Pierre Hayette; Nadine Cambisano; Nathalie Renotte; Olivier Ek; Sébastien Bontems; Vincent Bours |
| EPI_ISL_7440440, EPI_ISL_7442466 | SYNLAB MVZ Weiden | Robert Koch Institute | |
| EPI_ISL_7200823 | Salzkammergutklinikum Vöcklabruck, Institut für Pathologie | Salzkammergutklinikum Vöcklabruck, Institut für Pathologie | Franz Pühringer; Penka Lechner; Regina Stitz; René Silye; Senka Rohregger |
| EPI_ISL_7015235 | Selangor State Health Department | Institute for Medical Research, Infectious Disease Research Centre, National Institutes of Health, Ministry of Health Malaysia | Ahmad FA; Ahmad Fazilah NA; Anasir MI; Azizan MA; Kamel K; Mohamad Sukri MZ; Mohd Zawawi Z; Norhisham SN; Ramly N; Robert F; Rosli NR; Suppiah J; Thayan R |
| EPI_ISL_7467969 | Servicio Virosis Respiratorias-Departamento Virología-INEI | Instituto Nacional Enfermedades Infecciosas C.G.Malbran | Avaro M.; Baumeister E.; Benedetti E.; Campos J.; Cisterna D.; Dattero ME; De Belder D.; Haim MS.; Lorenzo F.; Molina V.; Perandones C.; Poklepovich T.; Pontoriero A.; Russo M.; Sanchez Loria J.; Tuduri E. |
| EPI_ISL_6795212, EPI_ISL_6825365, EPI_ISL_7056045, EPI_ISL_7056614 | Shamir Medical Center (Asaf Harofe) | Shamir Medical Center (Asaf Harofe) | Abu Hamad Ramzia; Adina Bar Chaim; Alona Frenkel; Anna Vishnevsky; Chen Weiner; Nir Rainy; Patricia Benveniste-Lekovitz; Reut Sorek Abramovich; Yevgeni Yegorov |
| EPI_ISL_7062525, EPI_ISL_7405371 | Spital Riggisberg | Institute for Infectious Diseases, University of Bern | Alban Ramette; Christian Baumann; Cora Sägesser; Franziska Suter-Riniker; Loïc Borcard; Miguel A Terrazos Miani; Nicole Liechti; Pascal Bittel; Peter Keller; Sonja Gempeler; Stefan Neuenschwander; Stephen L Leib |
| EPI_ISL_6963509 | Sri Jayadeva Institute of Cardiovascular Sciences and Research / Strand Life Sciences | National Centre for Biological Sciences, TIFR - Rockefeller Foundation | Aarati Karaba; Anson Kunjumon George; Aparnaa Ramanathan; Apurva Sarin; Chandrasekhar Vadlamudi; Chitra Pattabiraman; Darshan Sreenivas; Dasaradhi Palakodeti; Dimple Notani; Divya Priya A; Madhusudhan J; Manisha Bharadwaj; Manoj Kumar Jha; Mudasir Nazaar; Pradeep B P; Priyanka Ananta Mulay; Ramesh Hariharan; Rohan Pais; Satyajit Mayor; Saumitra Mardikar; Srivathsan Adimoolam; Uma Ramakrishnan; Vamsi Veeramachaneni; Vasanthapuram Ravi; Vijay Chandru; Vishal G Rao; Yasodha Kannan |
| EPI_ISL_7265233 | St Vincent's Pathology (SydPath) | NSW Health Pathology - Institute of Clinical Pathology and Medical Research; Westmead Hospital; University of Sydney | Arnott A.; Draper J.; Gall M.; Martinez E.; Rockett R.; Sintchenko V.; on behalf of ICPMR |
| EPI_ISL_7170972 | Stadtspital Triemli | Institute of Medical Virology | Alexandra Trkola; Annette Audigé; Cyril Shah; Gabriela Ziltener; Guido Bloemberg; Jon Huder; Jürg Böni; Kevin Steiner; Maria Grünberg; Maryam Zaheri; Michael Huber; Riccarda Capaul; Stefan Schmutz; Verena Kufner |
| EPI_ISL_6971472 | Stadtspital Triemli | Institute of Medical Virology, University of Zurich | Alexandra Trkola; Annette Audigé; Cyril Shah; Gabriela Ziltener; Guido Bloemberg; Jon Huder; Jürg Böni; Kevin Steiner; Maria Grünberg; Maryam Zaheri; Michael Huber; Riccarda Capaul; Stefan Schmutz; Verena Kufner |
| EPI_ISL_7445855 | State Hygienic Laboratory at the University of Iowa | State Hygienic Laboratory at the University of Iowa | Alankar Kampoowale; Anna Yakos; Cindy Toll; Davis Rieckenberg; Erik Twait; Jeff Benfer; Kris Eveland; Krishnaveni Sompallae; Kristen Zanon; Mariah Knutson; Mohammed Allam; Valerie Reeb; Wes Hottel |
| EPI_ISL_7013425, EPI_ISL_7264087, EPI_ISL_7264088, EPI_ISL_7456351, EPI_ISL_7456393, EPI_ISL_7456394, EPI_ISL_7456395, EPI_ISL_7456396, EPI_ISL_7456397, EPI_ISL_7456398, EPI_ISL_7456399, EPI_ISL_7456400 | | | |
| see above | State Laboratories Division, Hawaii State Department of Health | State Laboratories Division, Hawaii State Department of Health | Ayana Garnet; Daniel Strange; Drew Kuwazaki; Edward Desmond; Pamela O'Brien; Razvan Sultana |
| EPI_ISL_7544013, EPI_ISL_7544494, EPI_ISL_7544584, EPI_ISL_7544730 | Study: Rapid Diag POC Covid | NHLS/UCT | Arash Iranzadeh; Bruna Galvao; Carolyn Williamson; Deelan Doolabh; Diana Hardie; Gert Marais; Innocent Mudau; Luicer Olubayo; Lynn Tyers; Marvin Hsiao; Nokuzola Mbhele; Rageema Joseph; Stephen Korsman |
| EPI_ISL_7364700, EPI_ISL_7364701, EPI_ISL_7364702 | Summit Clinical Laboratories | City of Milwaukee Health Department Laboratory | Amy Bauer; Manjeet Khubbar; Samantha Scott; Sanjib Bhattacharyya |
| EPI_ISL_6883250, EPI_ISL_7452246, EPI_ISL_7452247 | Swedish national genomic surveillance program of SARS-CoV-2 | The Public Health Agency of Sweden | Alma Brolund; Maria Lind Karlberg; Maximilian Riess; Swedish national genomic surveillance program of SARS-CoV-2 |
| EPI_ISL_7457427, EPI_ISL_7457428, EPI_ISL_7457429, EPI_ISL_7457430, EPI_ISL_7457431 | Synlab Medilab | Karolinska University Hospital Huddinge | Annika Tiveljung Lindell; Henning Onsbring; Jan Albert; Karina Hentrich; Lynda Eneh; Martin Ekman; Natalija Gerasimcik; Robert Dyrdak; Sandra Broddesson; Shambhu Ganeshappa Aralaguppe; Tanja Normark; Tobias Allander; Valtteri Wirta; Zhibing Yun |

| | | | |
|---|---|---|---|
| EPI_ISL_7456440 | Tambo Memorial Laboratory | National Institute for Communicable Diseases of the National Health Laboratory Service | Amoako DG; Bhiman JN; Everatt J; Ismail A; Mahlangu B; Mnguni A; Mohale T; Ntuli N; Scheepers C; Wolter N |
| EPI_ISL_6825551 | Territory Pathology | Territory Pathology | Dimitrios Menouhos; Ella Meumann; Robert Baird |
| EPI_ISL_7171744 | The Hope Clinic of Emory Vaccine Center, Emory University | Piantadosi Lab, Emory Department of Pathology | Anne Piantadosi; Azmain Taz; Dara Khosravi; Ethan Wang; Jesse Waggoner; Ludy Carmola; Marybeth Sexton; Nadine Rouphael |
| EPI_ISL_7446810 | Thüringer Landesamtes für Verbraucherschutz | Robert Koch Institute | |
| EPI_ISL_7235629 | Tulane University School of Medicine | Tulane University School of Medicine | Di Tian |
| EPI_ISL_7505962 | UC Davis Genome Center | UC Davis Genome Center | Healthy Davis Together; UC Davis |
| EPI_ISL_7451261 | UNILABS | Instituto Nacional de Saude (INSA) | Borges et al |
| EPI_ISL_7160037, EPI_ISL_7160038, EPI_ISL_7160039, EPI_ISL_7545421, EPI_ISL_7545422, EPI_ISL_7545423, EPI_ISL_7545424 | | | |
| see above | UW Virology Lab | UW Virology Lab | Alexander Greninger; Hong Xie; Isabel Arnould; Keith R Jerome; Meei-Li Huang; Nathan Breit; Patrick Mathias; Pavitra Roychoudhury; Pooneh Hajian; Ricardo Perez; Robert J. Livingston; Saraswathi Sathees; Sean Ellis; Seffir T. Wendm; Shah Mohamed Bakhash; Tien V. Nguyen |
| EPI_ISL_7544861 | Unilabs | Institute of Medical Virology, University of Zurich | Alexandra Trkola; Annette Audigé; Cyril Shah; Gabriela Ziltener; Guido Bloemberg; Jon Huder; Jürg Böni; Kevin Steiner; Maria Grünberg; Maryam Zaheri; Michael Huber; Riccarda Capaul; Stefan Schmutz; Verena Kufner |
| EPI_ISL_7470330, EPI_ISL_7470341 | Unilabs Laboratory Medicine | Norwegian Institute of Public Health, Department of Virology | Atiya R Ali; Debech Nadia; Engebretsen Serina Beate; Garcia Llorente Ignacio; Hilde Elshaug; Hilde Vollan; Jon Bråte; Kamilla Heddeland Instefjord; Karoline Bragstad; Kathrine Stene-Johansen; Line Victoria Moen; Marie Paulsen Madsen; Olav Hungnes; Pedersen Benedikte Nevjen; Rasmus Riis Kopperud |
| EPI_ISL_7141056 | Unipath Speciality Laboratory Limited, Ahmedabad | Gujarat Biotechnology Research Centre | Apurvasinh Puvar; Bhadreshsinh Gohil; Chaitanya Joshi; Dinesh Kumar; Janvi Raval; Jwalant Shah; Madhvi Joshi; Nimesh Patel; Nitin Savaliya; Nitin Shukla; Priyank Chavda; Ramesh Pandit; Sonal Sharma; Zarna Patel |
| EPI_ISL_7542255 | University Medical Center Hamburg Eppendorf | Heinrich Pette Institute, Leibniz Institute for Experimental Virology | Adam Grundhoff; Alexis Robitaille; Johannes Knobloch; Martin Aepfelbacher; Nicole Fischer; Thomas Günther |
| EPI_ISL_6929785, EPI_ISL_7195724, EPI_ISL_7195725, EPI_ISL_7195726, EPI_ISL_7405404, EPI_ISL_7405405, EPI_ISL_7405406, EPI_ISL_7405407, EPI_ISL_7405408, EPI_ISL_7405409, EPI_ISL_7405412, EPI_ISL_7462253, EPI_ISL_7462258, EPI_ISL_7462265, EPI_ISL_7462271 | | | |
| see above | University of Liège COVID-19 testing center | GIGA Medical Genomics | Bouchra Boujemla; Claire Gourzonès; Cécile Meex; Keith Durkin; Laurent Gillet; Maria Artesi; Marie-Pierre Hayette; Nadine Cambisano; Nathalie Renotte; Olivier Ek; Sébastien Bontems; Vincent Bours |
| EPI_ISL_7379462 | University of Wisconsin-Madison AIDS Vaccine Research Laboratories | University of Wisconsin-Madison AIDS Vaccine Research Laboratories | Gage Moreno; Katarina Braun; et al. AIDS Vaccine Research Laboratories |
| EPI_ISL_7544862, EPI_ISL_7544872 | UniversitätsSpital Zürich | Institute of Medical Virology, University of Zurich | Alexandra Trkola; Annette Audigé; Cyril Shah; Gabriela Ziltener; Guido Bloemberg; Jon Huder; Jürg Böni; Kevin Steiner; Maria Grünberg; Maryam Zaheri; Michael Huber; Riccarda Capaul; Stefan Schmutz; Verena Kufner |
| EPI_ISL_7418452, EPI_ISL_7418453, EPI_ISL_7418454, EPI_ISL_7418455, EPI_ISL_7418456, EPI_ISL_7418463, EPI_ISL_7418464, EPI_ISL_7418465 | | | |
| see above | Universitätsklinikum Frankfurt - Institut für Medizinische Virologie | Robert Koch Institute | |
| EPI_ISL_7136300, EPI_ISL_7136771, EPI_ISL_7336152, EPI_ISL_7503247 | Utah Public Health Laboratory | Utah Public Health Laboratory | Erin L. Young; John Arnn; Kelly F. Oakeson; Olinto Linares-Perdomo; Pooja Gupta; Tom Iverson |
| EPI_ISL_7261603 | VA Tampa Healthcare System | VHA Public Health Reference Laboratory | Mark Holodniy on behalf of VA SEQFORCE; US Department of Veterans Affairs |
| EPI_ISL_6989662 | Vault Health | Minnesota Department of Health, Public Health Laboratory | Alyssa Mondelli; Elizabeth Horn; Jacob Garfin; Kelly Pung; Matt Plumb; Sarah Namugenyi; and Xiong Wang |
| EPI_ISL_7398681, EPI_ISL_7398758 | Vichaivej International Hospital Nongkhaem | National Institute of Health, Department of Medical Sciences, Ministry of Public Health, Thailand | Archawin Rojanawiwat; Ballang Uppapong; Beth Skaggs; Donlaya Maunplueng; Kazuhisa Okada; Natchaya Khiadsang; Nuttida Thongpramul; Pakorn Piromtong; Pilailuk Akkapaiboon Okada; Piroon Jenjaroenpun; Pongpun Sawatwong; Prapat Suriyaphol; Sirikanda Wimol; Siripaporn Phuygun; Sittiporn Parnmen; Supakit Sirilak; Suratchana Mitrat; Thanutsapa Thanadachakul; Thidathip Wongsurawat |
| EPI_ISL_7224567, EPI_ISL_7224577 | Viollier AG | Clinical Bacteriology, University Hospital Basel | Adrian Egli; Alfredo Mari; Christiane Beckmann; Fanny Wegner; Hans Hirsch; Helena MB Seth-Smith; Julia Bielicki; Karoline Leuzinger; Manuel Battegay; Tim Roloff |
| EPI_ISL_7371749, EPI_ISL_7371751, EPI_ISL_7372207 | Viollier AG | Department of Biosystems Science and Engineering, ETH Zürich | Andrea Patrignani; Andreia Cabral de Gouvea; Catharine Aquino; Chaoran Chen; Christiane Beckmann; Christoph Noppen; Daniel Ehrsam; Doris Popovic; Griffin White; Isabel Stürmer; Ivan Topolsky; Jay Tracy; Kim Philipp Jablonski; Lara Fuhrmann; Laura Neff; Lennart Opitz; Louis du Plessis; Maria Domenica Moccia; Maurice Redondo; Niko Beerenwinkel; Olivier Kobel; Ralph Schlapbach; Sarah Nadeau; Simon Grüter; Tanja Stadler; Timothy Sykes |
| EPI_ISL_7509012, EPI_ISL_7509019, EPI_ISL_7509025, EPI_ISL_7509030 | Virology Department, Royal Infirmary of Edinburgh, NHS Lothian / School of Biological Sciences, University of Edinburgh | COVID-19 Genomics UK (COG-UK) Consortium | Colquhoun R; Cotton S; Dewar R; Fernandez G; Gallagher A; Hill V; Jackson B; Maloney D; McCrone JT; McHugh M; O'Toole Á; Rambaut A; Scher E; Templeton K; Yu X |
| EPI_ISL_7285844, EPI_ISL_7285845, EPI_ISL_7285846, EPI_ISL_7285847, EPI_ISL_7285848 | Virology Laboratory, Scientific Department, Army Medical Center | Virology Laboratory, Scientific Department, Army Medical Center | Anella Monte; Anna Anselmo; Antonella Fortunato; Filippo Molinari; Florigio Lista; Francesco Giordani; Giancarlo Petralito; Giandomenico Cerreto; Giulia Campoli; Lucia Nicosia; Marzia Cavalli; Pietro Marco D'Angelo; Riccardo De Sanctis; Rossella Brandi; Silvia Fillo; Vanessa Vera Fain |
| EPI_ISL_7503508, EPI_ISL_7503509 | Washington State Department of Health Public Health Laboratories | Washington State Department of Health Public Health Laboratories | Alex Lathum; Ardizon Valdez; Avi Singh; Claire Howell; Denny Russell; Drew MacKellar; Holly Halstead; JohnAric Peterson; Kathryn Sickles; Kristin Roche; Lisa Jones; Philip Dykema; Rebecca Cao |
| EPI_ISL_7548944, EPI_ISL_7548945 | Wexner Medical Center | OSU College of Medicine | Corcoran, S.; Koenig, S. |
| EPI_ISL_7501188 | Winchester Hospital via Lahey Hospital | New England Biolabs | Abel, G.; B.W.; C.J.; Colgrove, R.; Duncan, R.; Elfahal, M.; Flynn; Heim, K.; Karolides, M.; L. and Langhorst; Michaels, L.; Pinet, K.; Skelton, T.; Sun |
| EPI_ISL_7263803 | Wisconsin State Laboratory of Hygiene Communicable Disease Division | Wisconsin State Laboratory of Hygiene Communicable Disease Division | Abigail C. Shockey; Alicia J. Mooney; Erika M. Hanson; Kelsey R. Florek; Richard Griesser; Sara Wagner; Tonya Danz |
| EPI_ISL_7478524, EPI_ISL_7478525, EPI_ISL_7478526, EPI_ISL_7478527, EPI_ISL_7478529, EPI_ISL_7478530, EPI_ISL_7502154, EPI_ISL_7502155 | | | |
| see above | Yale Clinical Virology Lab | Grubaugh Lab - Yale School of Public Health | Anderson Brito; Chaney Kalinich; Chantal Vogels; Isabel Ott; Joseph Fauver; Kendall Billig; Mallery Breban; Marie L. Landry; Mary Petrone; Nathan Grubaugh; Tobias Koch |
| EPI_ISL_7472293 | Yale Pathology Labs | Yale Pathology Labs | Angelique Levi; Brian Daley; Chen Liu; Guangxiao Yang; Heidi Herrick; Jianhui Wang; Jinglan Wang; John Sinard; Katherine Fajardo; Kevin Schofield; Laura Brady; Michael Stankewich; Minghao Zhong; Monica Talmor; Pei Hui; Peter Gershkovich; Richard Bouffard; Stephanie Weirsman; Susan Bell; Sylvia White |
| EPI_ISL_6795833, EPI_ISL_6795835, EPI_ISL_6795836, EPI_ISL_6795837, EPI_ISL_6795838, EPI_ISL_6795839, EPI_ISL_6795840, EPI_ISL_6795841, EPI_ISL_6795842, EPI_ISL_6795844, EPI_ISL_6795845, EPI_ISL_6795846, EPI_ISL_6795847, EPI_ISL_6795848, EPI_ISL_6795849, EPI_ISL_6795850, EPI_ISL_6825389, EPI_ISL_6825390, EPI_ISL_6825391, EPI_ISL_6825392, EPI_ISL_6825393, EPI_ISL_6825394, EPI_ISL_6825395, EPI_ISL_6825396, EPI_ISL_6825397, EPI_ISL_6825398, EPI_ISL_7015172, EPI_ISL_7015173, EPI_ISL_7015174, EPI_ISL_7015175, EPI_ISL_7015176, EPI_ISL_7015177, EPI_ISL_7015178, EPI_ISL_7015179, EPI_ISL_7015180, EPI_ISL_7015181, EPI_ISL_7015182, EPI_ISL_7015183, EPI_ISL_7015184, EPI_ISL_7015185, EPI_ISL_7015186, EPI_ISL_7015187, EPI_ISL_7015188, EPI_ISL_7015189, EPI_ISL_7015190, EPI_ISL_7015191, EPI_ISL_7015192, EPI_ISL_7015193, EPI_ISL_7015194, EPI_ISL_7015195, EPI_ISL_7015196, EPI_ISL_7015197, EPI_ISL_7015198, EPI_ISL_7015199, EPI_ISL_7015200, EPI_ISL_7015201, EPI_ISL_7015202, EPI_ISL_7015203, EPI_ISL_7015204, EPI_ISL_7015205, EPI_ISL_7015206, EPI_ISL_7015207, EPI_ISL_7015208 | | | |
| see above | ZARV/NHLS, Department Medical Virology, University of Pretoria | CERI, Centre for Epidemic Response and Innvoation, Stellenbosch University and KRISP, KZN Research Innovation and Sequencing Platform, UKZN. | Adriano Mendes; Amoaka D; Amy Strydom; Arisha Maharaj; Bester P; Bhiman J; Engelbrecht S; Everatt J; Giandhari J; Goedhals D; Hardie D; Hsiao M; Iranzadeh A; Lessells R; Makatini Z; Maponga T; Mdlalose N; Micheala Davids; Mlisana K; Moir M; NGS-SA (Scheepers C; Naidoo Y; Nyaga M) Giandhari J; Oluwakemi M; Pillay S; Preiser W; Ramphal U; Ramphal Y; San JE; Sim Mayaphi and Marietjie Venter; Tegally H; Tshiabuila D; Venter M; Wilkinson E; Williamson C; de Oliveira T; von Gottberg A |
| EPI_ISL_7548959, EPI_ISL_7548966, EPI_ISL_7549083 | ZOTZ KLIMAS MVZ Düsseldorf-Centrum GbR ÜBAG für Labormedizin, Genetik, Zytologie, Pathologie | Center of Medical Microbiology, Virology, and Hospital Hygiene, University of Duesseldorf | Alexander Dilthey; Andreas Walker; Daniel Strelow; Jessica Nicolai; Jörg Timm; Katrin Hoffmann; Klaus Pfeffer; Lisanna Hülse; Malte Kohns Vasconcelos; Maximilian Damagnez; Nadine Lübke; Patrick Finzer; Rainer Zotz; Tobias Wienemann; Torsten Houwaart |
| EPI_ISL_7544865 | Zentrallabor Zürich | Institute of Medical Virology, University of Zurich | Alexandra Trkola; Annette Audigé; Cyril Shah; Gabriela Ziltener; Guido Bloemberg; Jon Huder; Jürg Böni; Kevin Steiner; Maria Grünberg; Maryam Zaheri; Michael Huber; Riccarda Capaul; Stefan Schmutz; Verena Kufner |
| EPI_ISL_7423603 | amedes MVZ für Laboratoriumsdiagnostik Raubling GmbH | Robert Koch Institute | |
| EPI_ISL_7368223 | labor team w AG | Department of Biosystems Science and Engineering, ETH Zürich | Andrea Patrignani; Andreas Lindauer; Andreia Cabral de Gouvea; Catharine Aquino; Chaoran Chen; Daniel Ehrsam; Doris Popovic; Griffin White; Isabel Stürmer; Ivan Topolsky; Jay Tracy; Kim Philipp Jablonski; Lara Fuhrmann; Laura Neff; Lennart Opitz; Louis du Plessis; Maria Domenica Moccia; Monika Bucher; Niko Beerenwinkel; Ralph Schlapbach; Rebekka Pohl; Sarah Nadeau; Simon Grüter; Tanja Stadler; Timothy Sykes |

We gratefully acknowledge the following Authors from the Originating laboratories responsible for obtaining the specimens, as well as the Submitting laboratories where the genome data were generated and shared via GISAID, on which this research is based.

All Submitters of data may be contacted directly via www.gisaid.org

Authors are sorted alphabetically.

| Accession ID | Originating Laboratory | Submitting Laboratory | Authors |
|---|---|---|---|
| EPI_ISL_7190366 | PHV-FSS | PHV-FSS | Chenwei Wang on behalf of Q-PHIRE Genomics |
| EPI_ISL_7259544, EPI_ISL_7259701, EPI_ISL_7259710, EPI_ISL_7259716, EPI_ISL_7259721 | Public Health Ontario Laboratory | Public Health Ontario Laboratory | Aimin Li; Alireza Eshaghi; Andre Villegas; Ashleigh Sullivan; Christine Frantz; Dean Maxwell; Esha Joshi; Jared Simpson; Jennifer L Guthrie; Jonathan B Gubbay; Karthikeyan Sivaraman; Lawrence Heisler; Matthew Watson; Michael CY Li; Michael Laszloffy; Nahuel Fittipaldi; Philip Banh; Richard de Borja; Samir N Patel; Sandeep Nagra; Sandra Zittermann; Sarah Teatero; Vanessa G Allen; Yao Chen; Yogi Sundaravadanam |
| EPI_ISL_6795834 | ZARV/NHLS, Department Medical Virology, University of Pretoria | CERI, Centre for Epidemic Response and Innvoation, Stellenbosch University and KRISP, KZN Research Innovation and Sequencing Platform, UKZN. | Adriano Mendes; Amoaka D; Amy Strydom; Arisha Maharaj; Bester P; Bhiman J; Engelbrecht S; Everatt J; Goedhals D; Hardie D; Hsiao M; Iranzadeh A; Lessells R; Makatini Z; Maponga T; Mdlalose N; Micheala Davids; Mlisana K; Moir M; NGS-SA (Scheepers C; Naidoo Y; Nyaga M) Giandhari J; Oluwakemi M; Pillay S; Preiser W; Ramphal U; Ramphal Y; San JE; Sim Mayaphi and Marietjie Venter; Tegally H; Tshiabuila D; Venter M; Wilkinson E; Williamson C; de Oliveira T; von Gottberg A |
